# Source-water salinity measured by electrical conductivity in Bangladesh, 2025: a nationally representative cross-sectional survey

**DOI:** 10.64898/2026.10.10.26364870

**Authors:** S. M. Ashraf Ul Hoque, Md Khairul Islam, Mohammad Khalid Hasan, Md Ashifur Rahman, Saad Mohammad Quaish, Bilkis Banu, Zenat Zebin Hossain, Kamran ul Baset

## Abstract

**Background:** In coastal Bangladesh, salty drinking water has been linked to higher blood pressure. The 2025 Multiple Indicator Cluster Survey measured electrical conductivity, a marker of salinity, in household water sources. Its report did not estimate how many people were affected or where they lived.

**Objectives:** We estimated how many people live in households whose tested water source has high salinity and where they live. We also examined differences by household characteristics and source type, and overlap with arsenic.

**Methods:** Electrical conductivity was measured in the water source of 6,040 households in all 64 districts, February to June 2025. The main outcomes were conductivity above 1,500 and above 3,000 µS/cm, the survey’s risk classes, not health-based standards. Estimates used the survey weights and design.

**Results:** The tested source exceeded 1,500 µS/cm in 5.3% of households (95% confidence interval 4.7 to 5.9). Meter calibration was not documented, and most scenarios gave 4.5% to 6.2%. About 9.0 million people (7.9 to 10.1) lived in these households, including 1.8 million above 3,000 µS/cm. The 19 coastal districts held 26% of households but 70% of those above 1,500 µS/cm. Prevalence was similar across wealth groups and in households with a pregnant woman. Within the coastal zone, deep and shallow tube wells had high conductivity in similar shares (16.1% against 17.9%). After adjustment for district, deep wells had about 18% lower conductivity, but this difference was smaller and uncertain within subdistricts and differed by region.

**Conclusions:** Millions of people live in households whose water source is highly saline, mostly in the coastal zone. Deep wells were not reliably less saline than shallow wells, and a source that is safe for arsenic may still be salty. National water surveys should measure salinity alongside arsenic and test water in more than one season.

## Introduction

Salt water moving into coastal groundwater is projected to reach at least one kilometre inland in 41 countries by 2050 [1]. The seven most vulnerable low-and middle-income countries are all in South and Southeast Asia [1]. In coastal Bangladesh, seawater intrusion, cyclone surges and lower river flow from upstream have made drinking water salty, and rising sea levels are expected to make this worse [2,3]. About 20 million coastal residents are often described as affected, but this figure was carried forward from earlier secondary estimates [4,5]. Another estimate put more than 35 million coastal residents at risk as their fresh water turns salty [6]. Neither figure comes from measuring the water people use.

Salty drinking water adds sodium on top of the salt in food [2]. Dacope, a coastal subdistrict of Khulna, shows how large this can be. In the dry season, pregnant women there took an estimated 5 to 16 g of salt a day from drinking water, against about 1 g in the rainy season [2]. High blood pressure in pregnancy was more than twice as common in the dry season [2]. In a case-control study in coastal Bangladesh, the odds of pre-eclampsia or gestational hypertension were 5.5 times higher when water sodium was above 900 mg/L than below 300 mg/L [7]. In a cohort of 581 adults in coastal Bangladesh, lower water sodium was linked to lower blood pressure [8].

A systematic review rated this evidence as moderate [5]. A 2025 meta-analysis of 27 observational studies linked saltier water to higher systolic and diastolic blood pressure, and to 26% higher odds of hypertension [9]. Two earlier analyses did not show this pattern. A meta-analysis of seven mostly cross-sectional studies found no clear link with systolic blood pressure [10]. In pooled cohort data, mildly salty water was linked to lower blood pressure than fresh water, probably because it also carries calcium and magnesium [11]. The World Health Organization has set no health-based limit for sodium in drinking water; it gives 200 mg/L as the level above which taste becomes unacceptable [12].

Bangladesh has no national estimate of measured source-water salinity by source type and place [13]. Its 2009 national drinking-water quality survey measured chloride, one of the salts in seawater, in only 543 households and was not designed to give district estimates. Local studies show severe salinity in single coastal subdistricts [2,6], across nine coastal districts [4] and in aquifer sampling [14,15]. Salinity is usually measured as electrical conductivity (EC), which rises with the amount of salt dissolved in water. One survey of 30 south-west wells found a mean EC of 7,136 µS/cm [15]. These local studies cannot show how many people nationally use salty source water, where they live, which households they are, or whether deep tube wells are less salty. This matters because deep tube wells are the source promoted to avoid arsenic, and about three quarters of the south-west coastal population relied on them in 2015 [16].

Since 2014, Multiple Indicator Cluster Surveys (MICS) have tested household drinking water for *E. coli* with portable kits [17]. To our knowledge, the Bangladesh MICS 2025 is the first MICS round to measure EC in source water [18]. Its report gives risk classes by division, residence, wealth and tested source, but no district or coastal-zone estimates, no counts of people and no link with arsenic.

The overall objective was to describe exposure to high source-water EC across Bangladesh, using MICS 2025. Specifically, the study estimated how many households and people have a source with high EC, and where they live, including the coastal zone. It compared prevalence by household wealth, residence, the presence of young children or a pregnant woman, and how households obtained water. It also compared EC between deep and shallow tube wells, and estimated how many people live in households with both high EC and arsenic. Differences by month of testing were explored. The survey did not measure health, so this study describes exposure, not health effects. S1 Fig shows which steps from environment to health this study measured, which it estimated, and which rest on other evidence.

## Methods

### Data, sampling and measurement

This study is a secondary analysis of the Bangladesh MICS 2025, a nationally representative household survey run by the Bangladesh Bureau of Statistics and UNICEF Bangladesh [18]. The survey used a stratified two-stage cluster design: clusters were sampled first, then households within them. It interviewed 61,207 of 61,470 occupied households (99.6%). Data were collected on tablets by 43 field teams, each with five interviewers, one measurer and a supervisor. Measurers had 15 days of training in water testing, and supervisors re-interviewed one household per cluster. In each cluster, 2 of the 20 sampled households were chosen at random for water testing. The survey’s water-quality weight accounts for this subsampling and for households whose water was not tested. It was used as supplied, with the strata and clusters, for all estimates. S1 Table shows the sample flow. The survey set the sample size, so the precision of estimates is reported instead of statistical power.

The measurer rinsed a plastic cup, filled it with source water and recorded the EC reading and its unit. No meter model, calibration or temperature-correction procedure is documented. Readings recorded in millisiemens per centimetre (225 of 6,040) were converted to µS/cm. Fieldwork ran from 24 February to 30 June 2025. Two tests dated July were treated as recording errors and left out of the month analysis.

### Variables

#### Outcomes

EC measures all the salts dissolved in water. It indicates salinity but cannot show how much of it is sodium, which was not measured. The main outcomes were EC above 1,500 and above 3,000 µS/cm, the survey’s moderate and high risk classes [18]. These classes are not health-based standards. At 0.64 mg/L per µS/cm, they equal about 1,000 and 2,000 mg/L of total dissolved solids (TDS). Bangladesh’s Environment Conservation Rules of 1997 and 2023 set a TDS limit of 1,000 mg/L, which equals 1,560 µS/cm. Above this level, water tastes increasingly unpleasant [12], and coastal studies class it as slightly saline [4,16]. Secondary cut-offs were 700 µS/cm, the Food and Agriculture Organization fresh-water limit used in cohort studies [11], and 1,000 and 2,000 µS/cm. Approximate sodium values for coastal sodium-chloride water, from two published conversions, are used only to relate EC to health studies. They are least certain for deep wells, as 75% of deep aquifer samples in the south-west were sodium bicarbonate type [19].

Household prevalence is the percentage of households whose tested source was above a cut-off. Population prevalence is the percentage of people living in such households, weighting each household by its size. Population burden is the number of these people. It counts people whose household source was above the cut-off, not people whose own intake was measured.

#### Household characteristics

Residence, wealth quintile and household size came from the survey files. Wealth quintiles are based on the survey’s index of household assets and housing [18]. A child under 5 was identified from the household listing, and a current pregnancy from the interview with women aged 15 to 49. A main source counted as outside the dwelling or yard if it was located elsewhere, piped to a neighbour or public tap, or delivered. Water treatment and use of a different wet-season source were also described. Race and ethnicity were not analysed.

#### Place, source and arsenic

The coastal zone was the 19 districts of the Coastal Zone Policy 2005 [20]. Place was also described by division, district and upazila (subdistrict). The survey’s geospatial file gave cluster elevation and 2020 rainfall (Climate Hazards Group InfraRed Precipitation with Station data). The survey moved each cluster’s coordinates a short distance within its upazila, so mapped locations are approximate. District boundaries came from geoBoundaries [21]. Source type is the source the water sample came from (the tested source): shallow tube well, deep tube well, piped water or other. The survey does not define the depth that separates deep from shallow wells. The tested source was not always the main drinking-water source, so the unit of exposure is the tested source, not the water household members drink. Arsenic above 10 µg/L followed the survey definition and came from a field kit with an undocumented detection limit. The survey also recorded which measurer did each test. The data identify 41 measurers, two fewer than the 43 teams in the survey report; the files do not explain the difference.

### Statistical analysis

#### General approach

Analyses were primary (prevalence, population burden and geographic patterns), secondary (household characteristics, source type and arsenic), sensitivity or exploratory (month). The analysis was not pre-registered, and exploratory P values were not corrected for multiple testing. S2 Table lists each model with its reference groups and sample. No fixed significance threshold was used; estimates are interpreted through their 95% confidence intervals. Reporting follows the STROBE guideline for cross-sectional studies (S1 Checklist).

All estimates used the survey weights, strata and clusters, with Taylor linearisation. Analyses used R 4.6.1 (packages survey 4.5, lme4 2.0.6, mgcv 1.9.4 and spdep 1.4.2), and the main estimates were reproduced in Stata 17 (S3 Table). Prevalence is given with logit 95% confidence intervals. Prevalence ratios came from survey-weighted quasi-Poisson models with a log link.

#### Prevalence and place

Counts of people multiplied population prevalence by the 2022 census population adjusted by the post enumeration check: 169,828,911 nationally, with matching totals for each division [22]. For the 19 coastal districts, the 2022 census count of 43,818,348 [23] was multiplied by its division’s adjustment factor, giving 45.0 million. The other 45 districts make up the remaining 124.8 million. Counts for subgroups use their own census populations, so they do not sum exactly to the national count (Table 2). As a sensitivity analysis, counts were scaled to the middle of the testing period (factor 1.034). Prevalence ratios by residence and wealth quintile were adjusted for tested source and division. For how households obtained water, prevalence ratios compared a main source outside the dwelling or yard between coastal households above and at or below 1,500 µS/cm. These were adjusted for residence, wealth quintile and division, and then district. The coastal zone was compared with other districts by crude prevalence ratios, because source type lies on the path from place to EC. Moran’s I (eight nearest neighbours) measured whether nearby clusters had similar mean log10 EC. The map is a descriptive surface of cluster means and is not used for inference. District estimates are descriptive and are not ranked (S4 Table).

#### Source type

Deep and shallow tube wells were compared with survey-weighted linear models on log10 EC, which give geometric mean ratios (GMR). A GMR of 0.82 means that adjusted geometric mean EC was about 18% lower. Place affects both which source a household uses and how salty its groundwater is, so models were adjusted for factors that come before the choice of source. The declared primary model, fitted separately in the coastal zone and elsewhere, included district fixed effects, elevation, rainfall, residence and wealth. Crude, division, measurer and upazila models were sensitivity analyses (S2 Table). Zero readings (54) cannot be logged and were left out of these models. To test whether the coastal contrast differed by region, the primary model was refitted with an interaction between well type and three coastal regions. These were Khulna and Barishal divisions, Chattogram division, and Gopalganj and Shariatpur in Dhaka division; a design-based Wald test assessed the interaction.

#### Arsenic

Households were cross-classified by EC above 1,500 µS/cm and arsenic above 10 µg/L. People in each group were counted in the same way as for the other cut-offs.

#### Sensitivity and exploratory analyses

Supervisors tested 605 samples of bottled mineral water to check meter consistency. Readings of 20 µS/cm or less, far below the 5th percentile of 105 µS/cm, were treated as suspect. They were set to missing, counted as above each cut-off, or excluded. Other scenarios shifted or scaled all readings, bounded missing readings, kept only households whose tested source was their main source, and assumed an uncompensated meter (S3 Table). Unweighted mixed models estimated how much of the variation in EC lay between measurers. The month model added source type to the primary deep and shallow well model; it compares different samples tested in different months. As a check, six readings in Rangpur division were excluded. Each exceeded 3,000 µS/cm and was at least 9 times the reading of the other household in its cluster; they were identified by inspection (S3 Table). Other analyses used complete cases.

## Ethics approval and consent

MICS 2025 was approved by the icddr,b Ethical Review Committee (protocol PR-24113, November 2024), and verbal informed consent was obtained from all respondents [18]. This analysis used anonymised data supplied under the MICS data terms and needed no further approval. The data were accessed for research purposes on 26 September 2026. The authors had no access to information that could identify individual participants during or after data collection. The files contain no names or addresses, cluster coordinates were displaced by the survey, and no coordinates are reported in this paper.

## Results

Of 6,298 households selected for water testing, 6,111 (97.0%) completed the interview and 6,040 (98.8%) had an EC reading, from all 64 districts (S1 Table). Missing readings were more common in Khulna (4.4%) than in other divisions (0.3% to 1.4%; P < 0.001). They were not related to residence, wealth or household size (S1 Table). Of the 6,040 households, 69.8% were rural. The tested source was a shallow tube well in 55.4%, a deep tube well in 19.6% and piped water in 23.0% (Table 1). The 19 coastal-zone districts held 26.4% of weighted households (Table 1).

**Table 1.** Characteristics of the 6,040 households with an EC reading.

| Characteristic | Households (n) | Weighted % (95% CI) | Missing (n) |
| --- | --- | --- | --- |
| **Residence** |  |  | 0 |
| Urban | 1,637 | 30.2 (29.7 to 30.7) |  |
| Rural | 4,403 | 69.8 (69.3 to 70.3) |  |
| **Wealth quintile** |  |  | 0 |
| 1 (poorest) | 1,476 | 20.9 (19.8 to 22.1) |  |
| 2 | 1,296 | 20.0 (18.9 to 21.1) |  |
| 3 | 1,192 | 19.4 (18.3 to 20.6) |  |
| 4 | 1,098 | 19.8 (18.6 to 21.0) |  |
| 5 (richest) | 978 | 19.9 (18.8 to 21.2) |  |
| **Division** |  |  | 0 |
| Barishal | 535 | 5.3 (5.1 to 5.6) |  |
| Chattogram | 1,078 | 18.6 (18.1 to 19.0) |  |
| Dhaka | 1,366 | 26.7 (26.3 to 27.2) |  |
| Khulna | 872 | 11.6 (11.2 to 11.9) |  |
| Mymensingh | 358 | 7.6 (7.3 to 7.9) |  |
| Rajshahi | 735 | 13.7 (13.3 to 14.0) |  |
| Rangpur | 733 | 11.1 (10.8 to 11.4) |  |
| Sylhet | 363 | 5.4 (5.2 to 5.6) |  |
| **Zone** |  |  | 0 |
| Coastal zone (19 districts) | 1,771 | 26.4 (25.9 to 26.9) |  |
| Other districts (45) | 4,269 | 73.6 (73.1 to 74.1) |  |
| **Tested source** |  |  | 0 |
| Shallow tube well | 3,436 | 55.4 (54.1 to 56.7) |  |
| Deep tube well | 1,458 | 19.6 (18.6 to 20.7) |  |
| Piped water | 979 | 23.0 (21.9 to 24.1) |  |
| Other source | 167 | 2.0 (1.6 to 2.4) |  |
| **Tested source against main source** |  |  | 0 |
| Same as main source | 5,175 | 85.5 (84.3 to 86.6) |  |
| Different from main source | 865 | 14.5 (13.4 to 15.7) |  |
| **Month of testing, 2025** |  |  | 0 |
| February | 305 | 5.3 (4.4 to 6.2) |  |
| March | 1,762 | 29.4 (27.8 to 31.2) |  |
| April | 1,594 | 26.5 (24.8 to 28.1) |  |
| May | 1,734 | 29.1 (27.6 to 30.6) |  |
| June | 643 | 9.7 (8.7 to 10.9) |  |
| July | 2 | 0.1 (0.0 to 0.2) |  |
| **Child under 5 in household** |  |  | 0 |
| Child under 5 | 2,058 | 34.5 (33.1 to 35.9) |  |
| No child under 5 | 3,982 | 65.5 (64.1 to 66.9) |  |
| **Pregnant woman in household** |  |  | 897 |
| Currently pregnant woman | 284 | 5.5 (4.8 to 6.2) |  |
| No pregnant woman recorded | 4,859 | 94.5 (93.8 to 95.2) |  |
| **Location of main drinking-water source** |  |  | 16 |
| In dwelling or yard | 4,919 | 85.0 (84.0 to 86.0) |  |
| Not in dwelling or yard | 1,105 | 15.0 (14.0 to 16.0) |  |
| **Household size, members** |  |  | 0 |
| Median (interquartile range) | 6,040 | 4.0 (3.0 to 5.0) |  |
| **Cluster elevation, m** |  |  | 0 |
| Median (interquartile range) | 6,040 | 11.2 (7.7 to 17.5) |  |
| **Annual rainfall 2020, mm** |  |  | 0 |
| Median (interquartile range) | 6,040 | 2,337 (2,120 to 3,039) |  |

Percentages use the water-quality weight and *n* are unweighted; the 95% confidence intervals are logit intervals. Continuous variables are weighted medians. Elevation and rainfall are cluster-level covariates from the survey geospatial file. Location of the main source is “not in dwelling or yard” when the source is located elsewhere, piped to a neighbour or public tap, or delivered. Bottled and refill water are missing. Pregnancy status is missing for households with no interviewed woman aged 15 to 49.

**Table 2.** Households and people above source-water EC cut-offs, nationally and by zone, Bangladesh 2025.

| EC cut-off (µS/cm) | Status | Households above, % (95% CI) | People in households above, millions (95% CI) | Coastal zone, households above, % (95% CI) | Other districts, households above, % (95% CI) |
| --- | --- | --- | --- | --- | --- |
| 1,500 | Primary (survey class “moderate”) | 5.3 (4.7 to 5.9) | 9.0 (7.9 to 10.1) | 14.1 (12.4 to 16.1) | 2.1 (1.7 to 2.7) |
| 3,000 | Primary (survey class “high”) | 1.05 (0.79 to 1.40) | 1.8 (1.3 to 2.4) | 3.06 (2.22 to 4.19) | 0.33 (0.17 to 0.62) |
| 700 | Secondary (Food and Agriculture Organization fresh-water limit) | 28.1 (26.9 to 29.5) | 48.2 (45.9 to 50.6) | 48.6 (45.8 to 51.4) | 20.8 (19.4 to 22.3) |
| 1,000 | Secondary | 11.8 (10.9 to 12.7) | 20.1 (18.5 to 21.8) | 27.4 (25.0 to 29.9) | 6.2 (5.4 to 7.1) |
| 2,000 | Secondary | 2.8 (2.4 to 3.3) | 4.8 (4.1 to 5.7) | 7.8 (6.5 to 9.3) | 1.0 (0.7 to 1.5) |

EC ranged from 0 to 20,010 µS/cm, with a median of 462 µS/cm (S3 Table). The estimates reproduced the survey report’s published risk classes in every row compared (S4 Table). Bottled water used for quality control read a median of 228 µS/cm (S2 Fig). Of 6,040 source readings, 106 were 20 µS/cm or less, and five of the 41 measurers recorded 48.1% of them (S3 Table). Under most measurement, missing-data and temperature scenarios, 4.5% to 6.2% of households were above 1,500 µS/cm; extreme scenarios gave 3.4% to 9.2% (S3 Table). Measurers accounted for 18% of the variation in log10 EC in unweighted mixed models (S3 Table).

The tested source exceeded 1,500 µS/cm in 5.3% of households (95% CI 4.7 to 5.9). These households were home to about 9.0 million people (7.9 to 10.1 million; Table 2). About 1.8 million people (1.3 to 2.4) lived in households above 3,000 µS/cm, and about 20 million (18.5 to 21.8) above 1,000 µS/cm (Table 2). In 14.5% of households, the tested source was not the main drinking-water source (Table 1). In the other 85.5%, prevalence was also 5.3% (4.7 to 5.9; S3 Table). Fig 1 shows the share of people above any EC value. Most people (71.6%) lived in households at or below 700 µS/cm, against 51.5% in the coastal zone (S5 Table).

**Fig 1.**
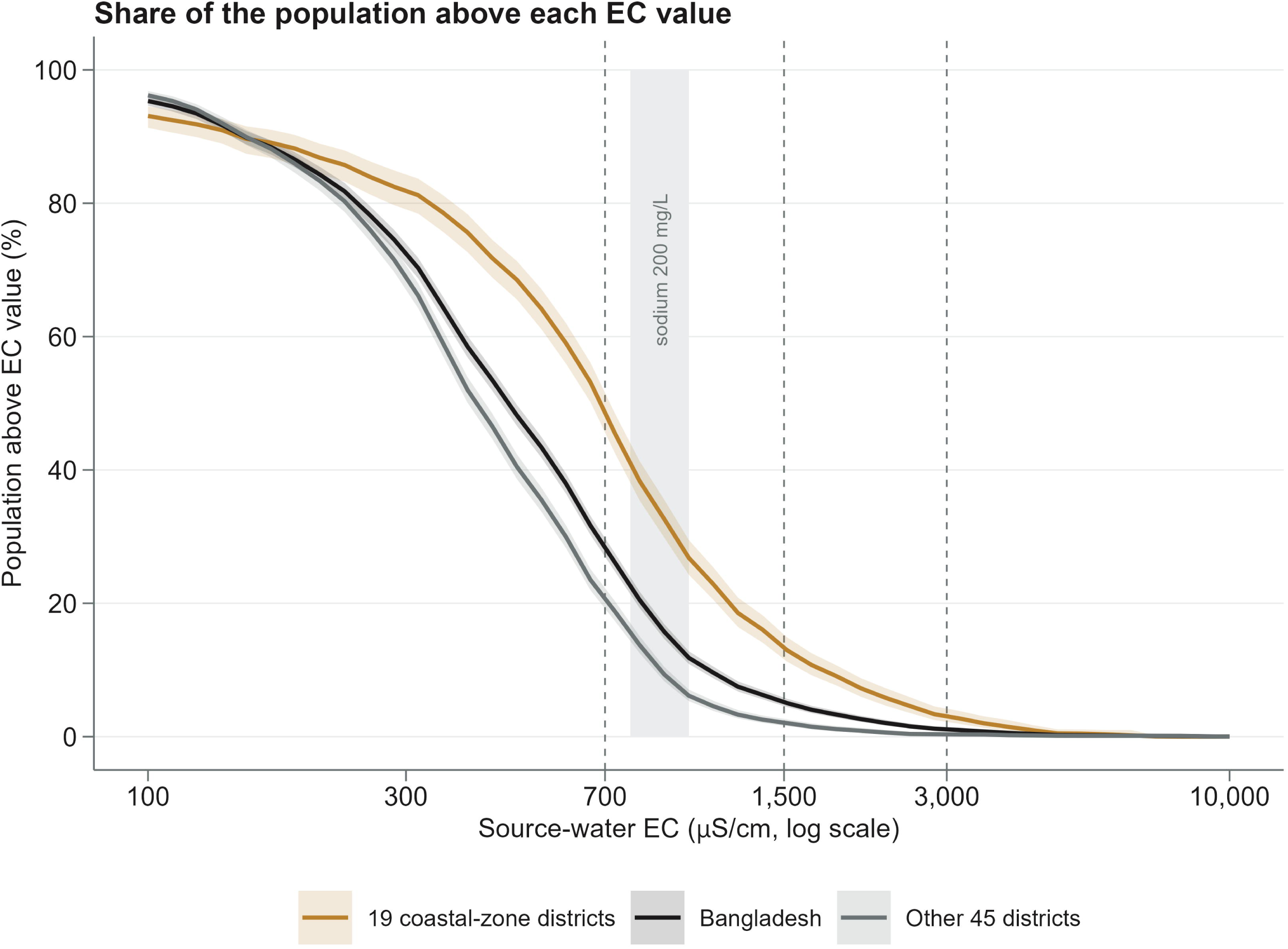
How many people live in households with high source-water EC. Survey-weighted share of the population in households whose tested source exceeded each EC value (95% pointwise confidence bands). Curves: Bangladesh, the 19 coastal-zone districts and the other 45 districts. Dashed lines: 700, 1,500 and 3,000 µS/cm. Grey band: EC at which sodium would be about 200 mg/L in coastal sodium-chloride water (780 to 1,000 µS/cm). Sensitivity of the 1,500 µS/cm estimate is in S3 Table. Data: Bangladesh MICS 2025; estimates use the survey weights and design.

Barishal (15.0%) and Khulna (14.5%) had the highest shares of households above 1,500 µS/cm, followed by Chattogram (7.7%) and Dhaka (3.7%). The other divisions had 1.7% or less (Fig 2C). By number of people, Chattogram and Khulna each had about 2.6 million, Dhaka 1.8 million and Barishal 1.3 million (Fig 2D). The coastal zone held 26.4% of households but 70.4% of those above 1,500 µS/cm (S5 Table). Its prevalence was 14.1%, against 2.1% in other districts (prevalence ratio 6.6, 95% CI 5.1 to 8.7; Table 2). Nearby clusters had similar EC (Moran’s I 0.585, P = 0.001; Fig 2A). Median cluster EC fell from 819 µS/cm at 5 m elevation or less to 188 µS/cm above 50 m (Fig 2B). With a median of 91 households per district, district estimates were imprecise (S4 Table). Point estimates exceeded 10% in 15 districts: 11 in the coastal zone and four inland (Faridpur, Madaripur, Meherpur and Munshiganj; S4 Table). Five clusters in Panchagarh, in the far north-west, had a mean above 3,000 µS/cm (Fig 2A). In each, one household read 8,105 to 9,906 µS/cm and the other 102 to 781 µS/cm, all recorded by the same measurer. This pattern suggests recording errors. A sixth discordant reading came from Gaibandha (3,338 µS/cm, against 370 µS/cm in the same cluster). Excluding all six changed the share of households above 3,000 µS/cm from 1.05% to 1.00% (S3 Table).

**Fig 2.**
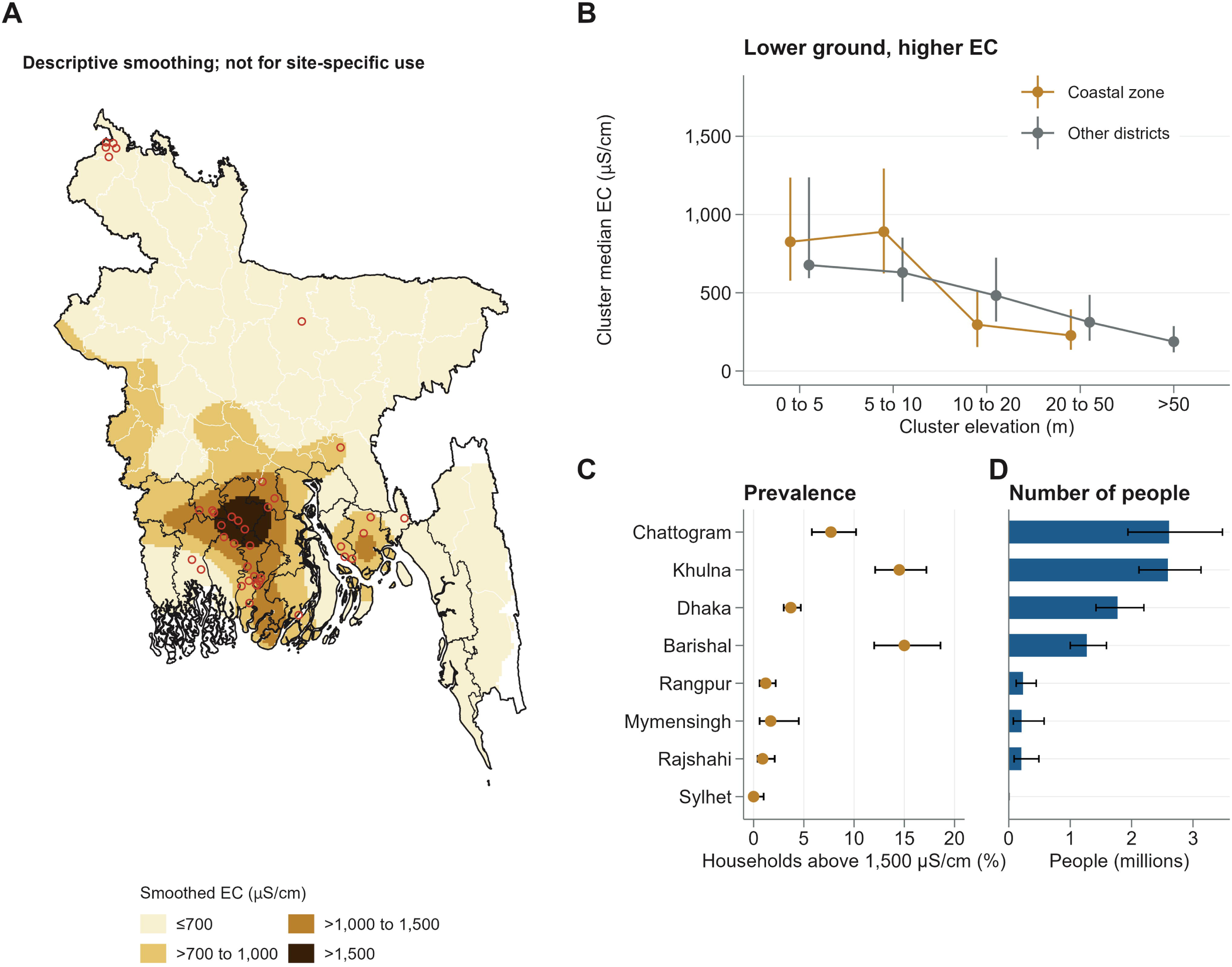
Where high source-water EC occurs. (A) Interpolated descriptive surface of cluster-mean EC (thin plate spline, log10 scale, 3 km grid within 15 km of a surveyed cluster); not a prediction at any location. Black lines mark the coastal-zone districts, and open red circles clusters with a mean above 3,000 µS/cm. Coordinates are displaced within upazila; boundaries from geoBoundaries [21]. (B) Median cluster EC (interquartile range) by elevation band; bands with fewer than 10 clusters omitted. (C) Household prevalence above 1,500 µS/cm by division (95% CI). (D) People living in households above 1,500 µS/cm by division (millions, 95% CI). Data: Bangladesh MICS 2025; estimates use the survey weights and design.

There was little evidence that high EC was linked to household wealth or urban residence. The adjusted prevalence ratio per wealth quintile was 1.02 (0.93 to 1.11) nationally and 0.99 (0.90 to 1.09) in the coastal zone (S5 Table). The adjusted prevalence ratio for urban against rural households was 0.90 (0.66 to 1.23; S5 Table). Households with a child under 5 had 5.3% (4.3 to 6.5) above 1,500 µS/cm. Households with a pregnant woman had 5.5% (3.5 to 8.5), an estimate based on only 284 households (Fig 3A). In the coastal zone, 44.5% of households above 1,500 µS/cm had their main source outside the dwelling or yard, against 31.6% of the others (Fig 3B). The adjusted prevalence ratio was 1.27 (1.08 to 1.49), and 1.19 (0.99 to 1.43) with district adjustment (S5 Table). Few coastal households treated their water (9.8%) or used a different wet-season source (S5 Table).

**Fig 3.**
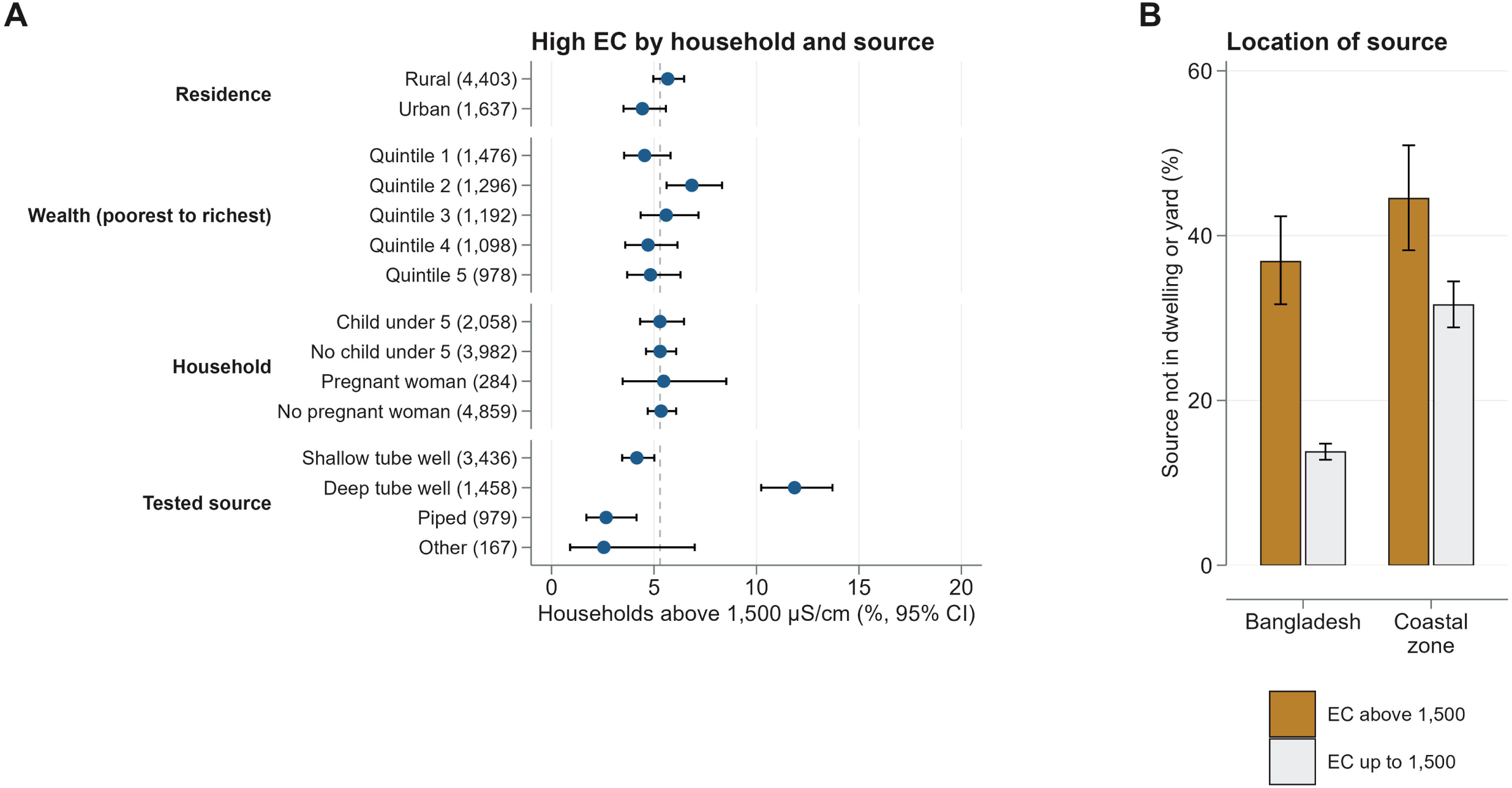
Which households have high source-water EC. (A) Household prevalence of EC above 1,500 µS/cm (95% CI) by residence, wealth quintile, child under 5, pregnant woman and tested source. The number of households is in brackets; the dashed line is the national prevalence (5.3%). Zone estimates are in Table 2. Source-type rows reflect where each source type is sited (60% of tested deep tube wells are in the coastal zone). (B) Share of households whose main source is not in the dwelling or yard, by EC above or up to 1,500 µS/cm (95% CI). Pregnancy status is available for 5,143 households. Data: Bangladesh MICS 2025; estimates use the survey weights and design.

Nationally, deep tube wells were above 1,500 µS/cm more often than shallow ones: 11.9% (10.2 to 13.7) against 4.2% (3.4 to 5.0; Fig 3A). Of tested deep wells, 59.9% were in the coastal zone, against 14.5% of shallow wells (S5 Table). The prevalence ratio was 2.85 (2.24 to 3.62) without adjustment and 0.79 (0.64 to 0.99) with district adjustment (S5 Table). In the coastal zone, 16.1% of deep and 17.9% of shallow wells were above 1,500 µS/cm (difference-1.7 percentage points, 95% CI-6.5 to 3.0; Table 3). Coastal deep wells more often had neither high EC nor arsenic above 10 µg/L: 79.1% against 47.8% of shallow wells (Table 3).

**Table 3.**
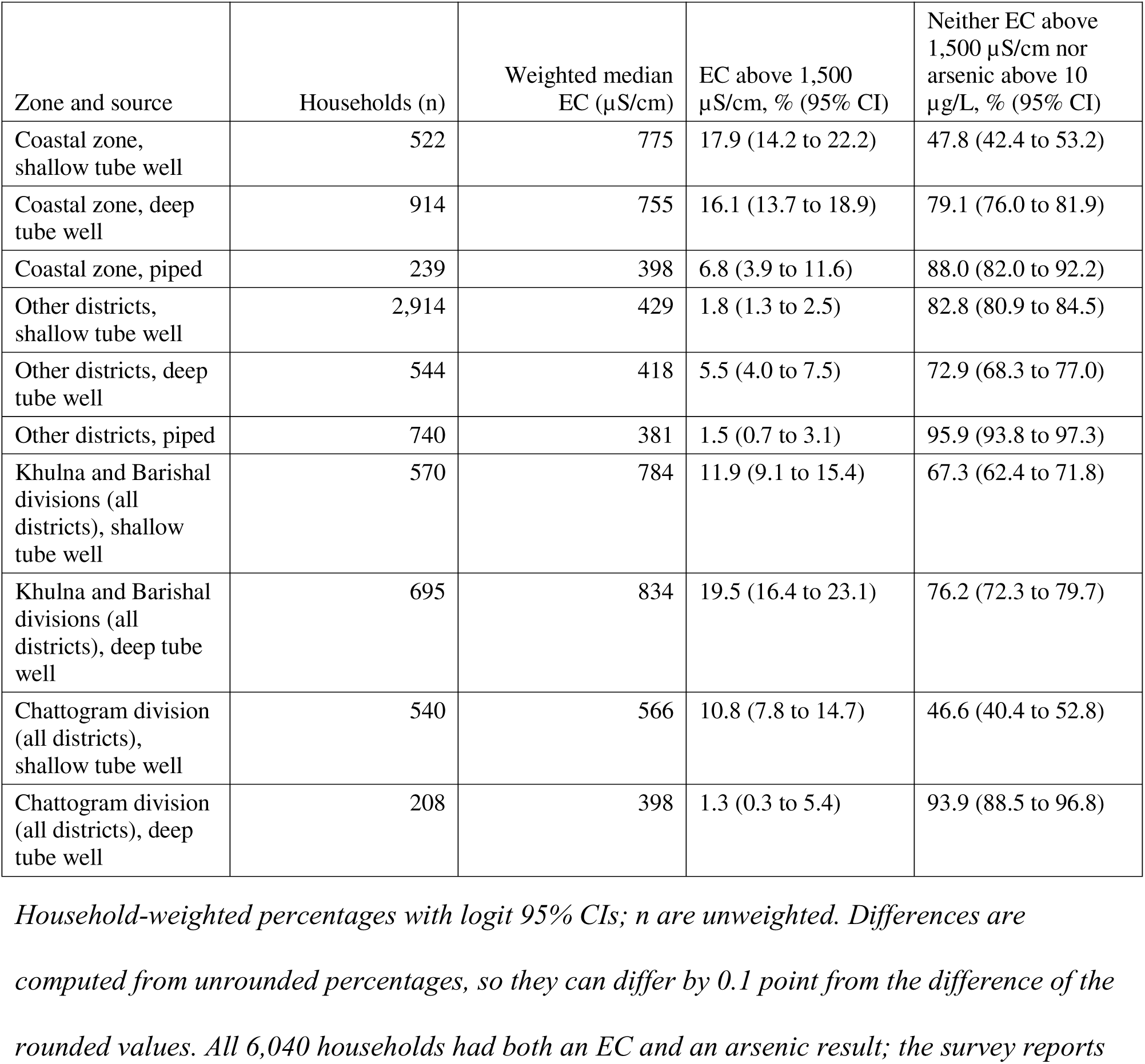
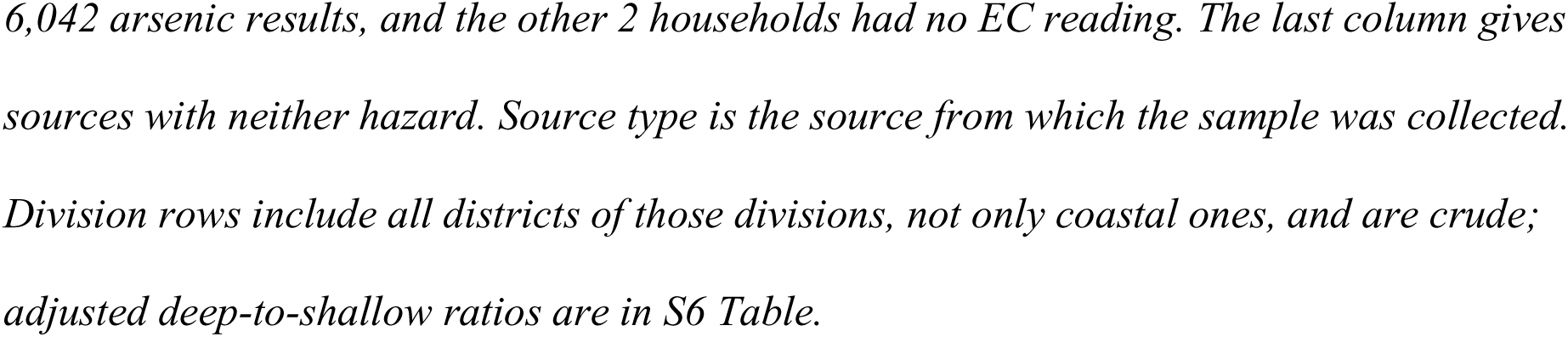
EC by tested source type, and sources with neither high EC nor arsenic, inside and outside the coastal zone (descriptive).

| Zone and source | Households (n) | Weighted median EC ($\mu\text{S}/\text{cm}$) | EC above 1,500 $\mu\text{S}/\text{cm}$, % (95% CI) | Neither EC above 1,500 $\mu\text{S}/\text{cm}$ nor arsenic above 10 $\mu\text{g}/\text{L}$, % (95% CI) |
| --- | --- | --- | --- | --- |
| Coastal zone, shallow tube well | 522 | 775 | 17.9 (14.2 to 22.2) | 47.8 (42.4 to 53.2) |
| Coastal zone, deep tube well | 914 | 755 | 16.1 (13.7 to 18.9) | 79.1 (76.0 to 81.9) |
| Coastal zone, piped | 239 | 398 | 6.8 (3.9 to 11.6) | 88.0 (82.0 to 92.2) |
| Other districts, shallow tube well | 2,914 | 429 | 1.8 (1.3 to 2.5) | 82.8 (80.9 to 84.5) |
| Other districts, deep tube well | 544 | 418 | 5.5 (4.0 to 7.5) | 72.9 (68.3 to 77.0) |
| Other districts, piped | 740 | 381 | 1.5 (0.7 to 3.1) | 95.9 (93.8 to 97.3) |
| Khulna and Barishal divisions (all districts), shallow tube well | 570 | 784 | 11.9 (9.1 to 15.4) | 67.3 (62.4 to 71.8) |
| Khulna and Barishal divisions (all districts), deep tube well | 695 | 834 | 19.5 (16.4 to 23.1) | 76.2 (72.3 to 79.7) |
| Chattogram division (all districts), shallow tube well | 540 | 566 | 10.8 (7.8 to 14.7) | 46.6 (40.4 to 52.8) |
| Chattogram division (all districts), deep tube well | 208 | 398 | 1.3 (0.3 to 5.4) | 93.9 (88.5 to 96.8) |

*Household-weighted percentages with logit 95% CIs; n are unweighted. Differences are computed from unrounded percentages, so they can differ by 0.1 point from the difference of the rounded values. All 6,040 households had both an EC and an arsenic result; the survey reports*
6,042 arsenic results, and the other 2 households had no EC reading. The last column gives sources with neither hazard. Source type is the source from which the sample was collected. Division rows include all districts of those divisions, not only coastal ones, and are crude; adjusted deep-to-shallow ratios are in S6 Table.

In the coastal zone, the GMR of deep against shallow wells was 1.09 (0.96 to 1.23) without adjustment and 0.82 (0.73 to 0.92) in the primary model (Fig 4B). Geometric mean EC was thus about 18% lower in sampled deep wells. This pooled ratio hid regional differences (interaction P < 0.001). The ratio was 0.70 (0.59 to 0.83) in coastal Chattogram, 0.86 (0.70 to 1.06) in Khulna and Barishal, and 1.43 (1.17 to 1.75) in Gopalganj and Shariatpur (S6 Table). In the 69 upazilas with both well types (851 households), the ratio was 0.93 (0.80 to 1.07; S6 Table). Excluding readings of 20 µS/cm or less gave 0.78 (0.72 to 0.86) with district adjustment and 0.88 (0.80 to 0.97) within upazilas (S6 Table). Outside the coastal zone, the primary ratio was 1.04 (0.96 to 1.13; S6 Table). In Khulna and Barishal combined, weighted mean TDS equivalents were 619 mg/L in shallow and 744 mg/L in deep tube wells (S5 Table).

**Fig 4.**
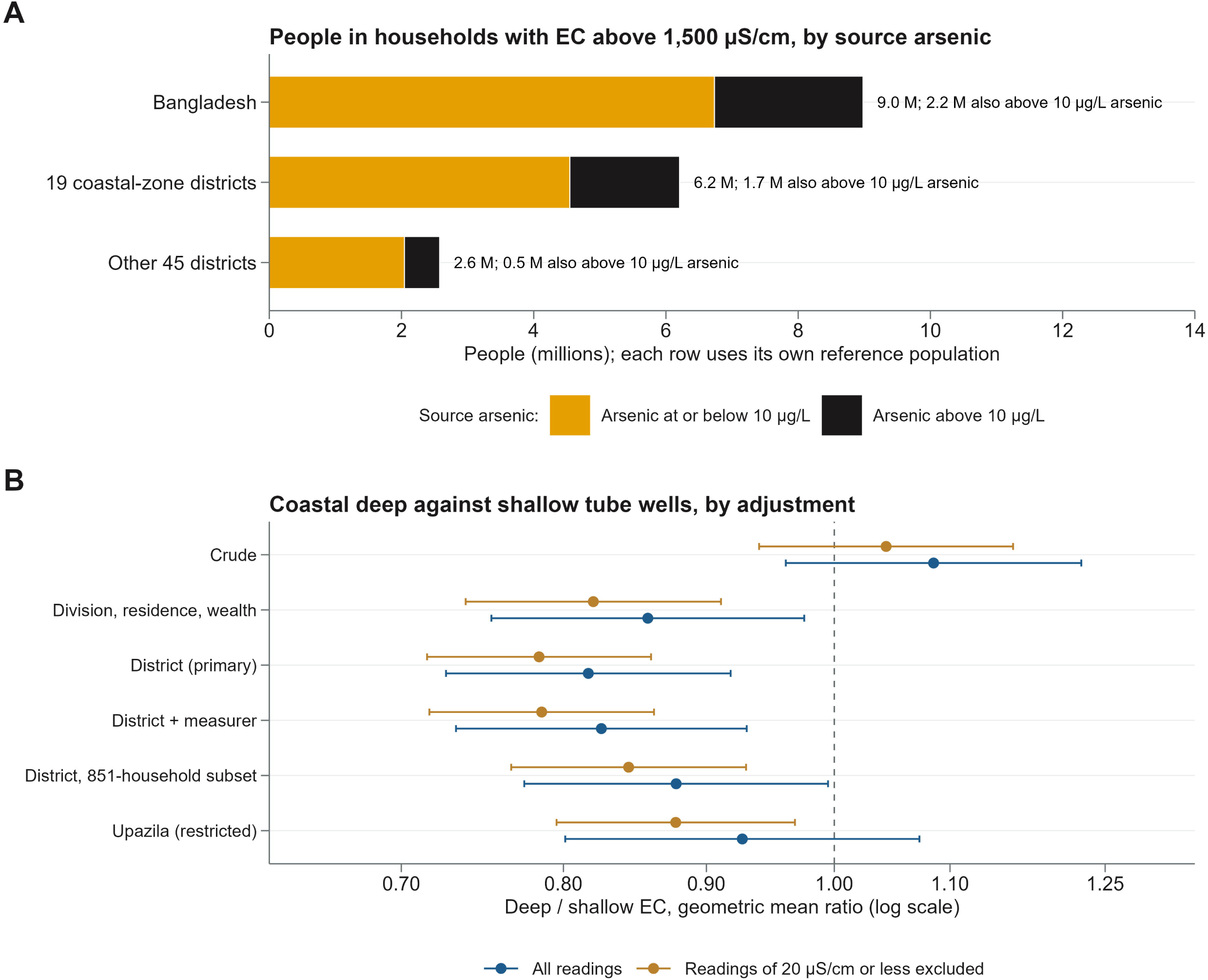
High EC with arsenic, and the deep against shallow contrast. (A) People (millions) living in households whose tested source had EC above 1,500 µS/cm, split by whether arsenic also exceeded 10 µg/L. Subgroup rows use their own reference populations and are not additive. (B) Geometric mean ratio of EC in deep against shallow coastal tube wells under each adjustment set (95% CI). Blue: all readings; gold: readings of 20 µS/cm or less excluded. District (primary) is the declared primary specification. The subset and upazila rows use the upazilas with both well types (851 households; 845 after exclusion). Ratios are conditional associations, not effects of well depth. Data: Bangladesh MICS 2025; estimates use the survey weights and design.

Of the 9.0 million people in households above 1,500 µS/cm, 2.2 million lived in households whose source also exceeded 10 µg/L arsenic (Fig 4A). Among people whose source had arsenic at or below 10 µg/L, 4.7% (4.1 to 5.3) lived in households above 1,500 µS/cm. In the coastal zone, this share was 12.3% (10.5 to 14.4; S5 Table).

In the coastal zone, weighted median EC rose from 594 µS/cm in February to 777 µS/cm in June (S5 Table). The adjusted ratio of May to June against February to March was 1.17 (0.98 to 1.39), and 0.86 to 1.30 in other specifications (S6 Table, Panel B). Khulna division supplied 15% to 17% of coastal tests above zero from February to April, but 33% to 41% in May and June (S5 Table).

## Discussion

In 2025, an estimated 9.0 million people in Bangladesh lived in households whose tested water source was above the survey’s moderate EC class. These included 1.8 million above the high class. The counts describe the tested source; they do not measure what people drink or any effect on health. As local studies of the coast and south-west suggested [4,15], these people were concentrated in the coastal zone, although a few inland districts also had high shares. The figures of 20 million and 35 million often quoted for the coast [4,5,6] did not come from measured water, so they cannot be compared directly with this count. Prevalence was similar across household wealth and in households with young children or pregnant women.

Nationally, deep tube wells had high EC more often than shallow wells, mainly because they are concentrated in salty coastal districts, where they were installed to avoid arsenic [16]. Within districts, deep wells had somewhat lower EC. This is an association, not evidence that depth itself lowers salinity. The difference also shrank within upazilas and changed with how very low readings were handled. A source with arsenic at or below 10 µg/L was not necessarily low in salinity.

Earlier national data allow only a rough comparison. In the 2009 survey, 2% of laboratory samples were above 600 mg/L chloride [13], which is roughly 1,500 µS/cm in sodium-chloride water. The share above that level in MICS 2025 was more than twice as high, but the two surveys differ in sampling, sample type and the salts measured.

Local studies report much higher exposure than this national picture. In one coastal subdistrict, 65% of young adults drank water above 600 mg/L salinity [6], and wells in a south-west survey averaged 7,136 µS/cm [15]. Such studies sample known hotspots, whereas a national random sample shows how far high EC extends beyond them. Seasonal rises in salt intake and blood pressure have been reported [2,4], and cyclone surges can make ponds and shallow groundwater salty within a short time [3]. A single dry-season survey may therefore miss short-lived peaks.

High EC followed place more than household wealth. Households with a pregnant woman were affected as often as others, and much of the health evidence comes from studies of pregnant women [2,7]. In the coastal zone, households with high-EC sources tended to fetch water from outside the dwelling or yard more often (district-adjusted prevalence ratio 1.19, 95% CI 0.99 to 1.43). Salty water and the work of collecting water may therefore affect the same households.

Studies of well depth give mixed results. A survey of nine coastal districts found higher mean TDS in deep (923 mg/L) than in shallow wells (837 mg/L) [4], the same order as in Khulna and Barishal here. In eight south-west districts, a third of deep tube-well samples were slightly or moderately saline [16]. Two smaller surveys found shallow wells saltier than deep wells [15,24]. In the south-west, median TDS was below 1,000 mg/L in the deep aquifer but far higher in the shallow aquifers [19]. In 46 deep wells in south-central Bangladesh, salinity did not rise between 1998 and 2011 [25]. The within-upazila estimate here was uncertain and does not show that deep and shallow wells are equally salty. The contrast also differed by region: deep wells were less saline than shallow wells in coastal Chattogram but more saline in Gopalganj and Shariatpur. A single pooled ratio therefore hides local differences in aquifers.

These EC levels can be related to sodium only roughly, and only in coastal sodium-chloride water. There, 1,500 and 3,000 µS/cm correspond to about 300 to 380 and 600 to 770 mg/L sodium. These levels are below the highest sodium groups in some studies (above 900 or 1,000 mg/L), but within lower groups in which risks were also raised. In a case-control study, the odds of pre-eclampsia or gestational hypertension were 3.3 times higher at 300 to 600 mg/L than below 300 mg/L [7]. Cohort studies have also linked water sodium to higher blood pressure [8,26], and diastolic pressure was raised above 1,000 mg/L [27]. In young adults in Koyra, a coastal subdistrict of Khulna, sodium above 600 mg/L was linked to systolic pressure higher by 3.5 mmHg [6]. Pooled over 27 studies, links with blood pressure were stronger in coastal populations [9]. EC cannot separate sodium from other salts, so these values are only a guide. S3 Fig places previous studies on the EC scale.

### Implications for water-quality surveillance

These points are the authors’ reading of the findings; this study did not test them. First, the results support measuring EC alongside arsenic in national and local water-quality surveys, building on the existing survey water-quality module [17]. This matters most in coastal districts and when new deep wells are sited. Deep wells are promoted because their arsenic is low [25], yet their EC was not reliably low. The survey’s current indicator, set at 3,000 µS/cm [18], captures only a small part of the population above the moderate class. Second, future MICS rounds should record the meter model, its calibration and temperature correction [12]. A laboratory subsample measuring sodium, calcium and magnesium would help, because minerals other than sodium may affect blood pressure [11]. A wet-season sample would show changes with season and after cyclones [2,3].

Third, other water options should be judged by their full mineral content, not by EC alone. Water from managed aquifer recharge and rainwater have both been linked to higher blood pressure in coastal Bangladesh [28,29]. Fourth, the Environment Conservation Rules 2023 replaced those of 1997 [30] and keep the drinking-water limits of 200 mg/L for sodium and 1,000 mg/L for TDS [31]. These equal the World Health Organization acceptability values, which are based on taste. Whether Bangladesh needs a health-based limit depends on health evidence that this study does not provide. One route is to link water-quality data with blood pressure in national health surveys, as done in India [32]. India has also mapped groundwater EC nationally and estimated that about 330 million people may be exposed above 2,000 µS/cm [33].

### Strengths and limitations

To our knowledge, this is the first analysis of a national random sample with measured source-water EC. It reproduced the survey report’s published risk classes in every row compared. Five limitations should guide how the findings are used. First, EC measures all dissolved salts, not sodium, so it cannot show who is harmed. Second, the tested source was not the main drinking-water source in 14.5% of households, and stored water was not measured. Prevalence was unchanged in households whose tested source was the main source, so this mismatch is unlikely to shift the national share. Testing covered only the dry and pre-monsoon season, so exposure over the whole year may be lower.

Third, the meter, its calibration and temperature correction were not documented. The scenarios cover random error and moderate bias, but a larger calibration error of unknown direction cannot be ruled out. Very low readings came mostly from a few measurers. This suggests meter or recording faults, although rainwater can truly read very low. The well-depth and month results changed when these readings were excluded. Arsenic came from a field kit, not a laboratory. Fourth, district and high-class estimates are imprecise, so districts are not ranked. Missing readings were most common in Khulna, a high-EC division, so the national share may be slightly underestimated. Counts based on the 2022 census are also slightly conservative. Fifth, the source-type and month comparisons are observational, and month is confounded with place and testing team. Local geology and actual well depth were not measured. Some wells recorded as deep may draw from shallower, saltier layers, which would hide part of any true difference by depth. Deep fresh water is also patchy: imaging along the Pusur River found two deep freshwater bodies separated by a high-salinity zone [34]. These comparisons therefore cannot show cause. The estimates apply to the tested sources of Bangladeshi households in the dry and pre-monsoon season of 2025.

## Conclusions

An estimated 9 million people in Bangladesh live in households whose tested water source exceeds 1,500 µS/cm, mostly in the coastal zone. Prevalence was similar across household wealth and in households with young children or pregnant women. Deep tube wells had somewhat lower EC after adjustment for place, but the difference shrank within upazilas, differed by region and cannot be attributed to depth. A source that was safe for arsenic was not necessarily low in salt. National water-quality surveys should therefore measure both, record meter calibration and test water in more than one season.

## Data Availability

MICS 2025 was approved by the icddr,b Ethical Review Committee (protocol PR-24113, November 2024), and verbal informed consent was obtained from all respondents. This analysis used anonymised data supplied under the MICS data terms and needed no further approval. The data were accessed for research purposes on September 2026. The authors had no access to information that could identify individual participants during or after data collection. The files contain no names or addresses, cluster coordinates were displaced by the survey, and no coordinates are shared.

https://mics.unicef.org

## Acknowledgments

The authors thank the UNICEF MICS programme and the Bangladesh Bureau of Statistics for the survey data.

## Supporting information

**S1 Checklist. STROBE checklist for cross-sectional studies.** (DOCX)

**S1 Table. Sample flow, non-measurement and analysis samples.** (DOCX)

**S2 Table. Model specifications.** (DOCX)

**S3 Table. Sensitivity of the main estimates to measurement, missing data and measurer.** (DOCX)

**S4 Table. Estimates by division, zone and district.** (DOCX)

**S5 Table. Numeric data for the figures and text.** (DOCX)

**S6 Table. Sensitivity of the depth and month comparisons to the specification.** (DOCX)

---

**S1 Fig.**
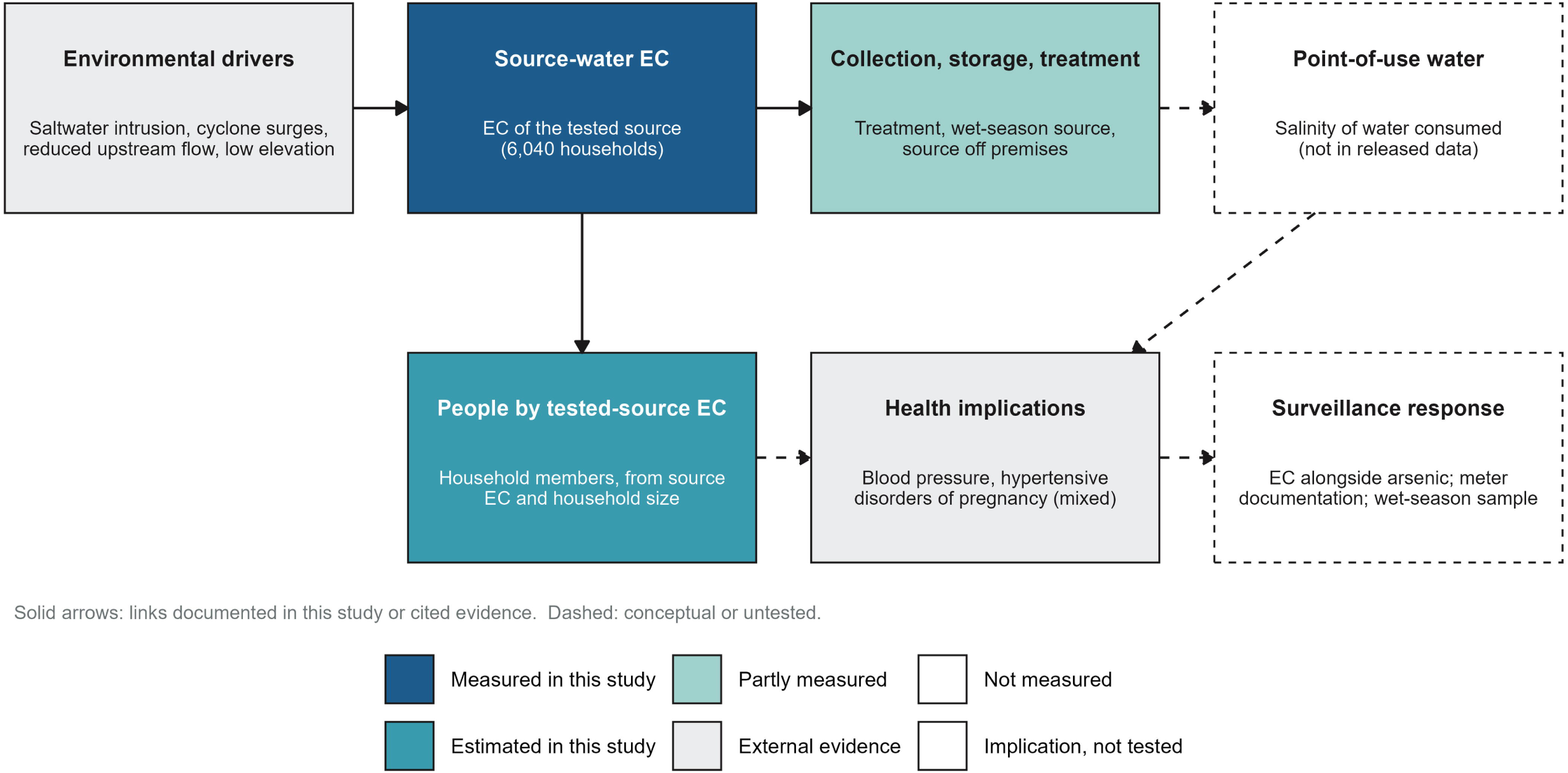
Conceptual framework for drinking-water salinity surveillance. Each box is labelled by its evidential status. Measured in this study: source-water EC. Estimated in this study: people by tested-source EC, from source EC and household size. Partly measured: household handling (treatment, wet-season source, collection off premises). Not measured: salinity at the point of use, which is not in the released data. External evidence: environmental drivers (main text references 2 to 4) and health implications, with mixed findings. (main text references 5, 7, 8, 10, 11, 26 and 27) Implication not tested: surveillance response. Dashed borders and arrows mark conceptual or untested links. (TIF)

**S2 Fig.**
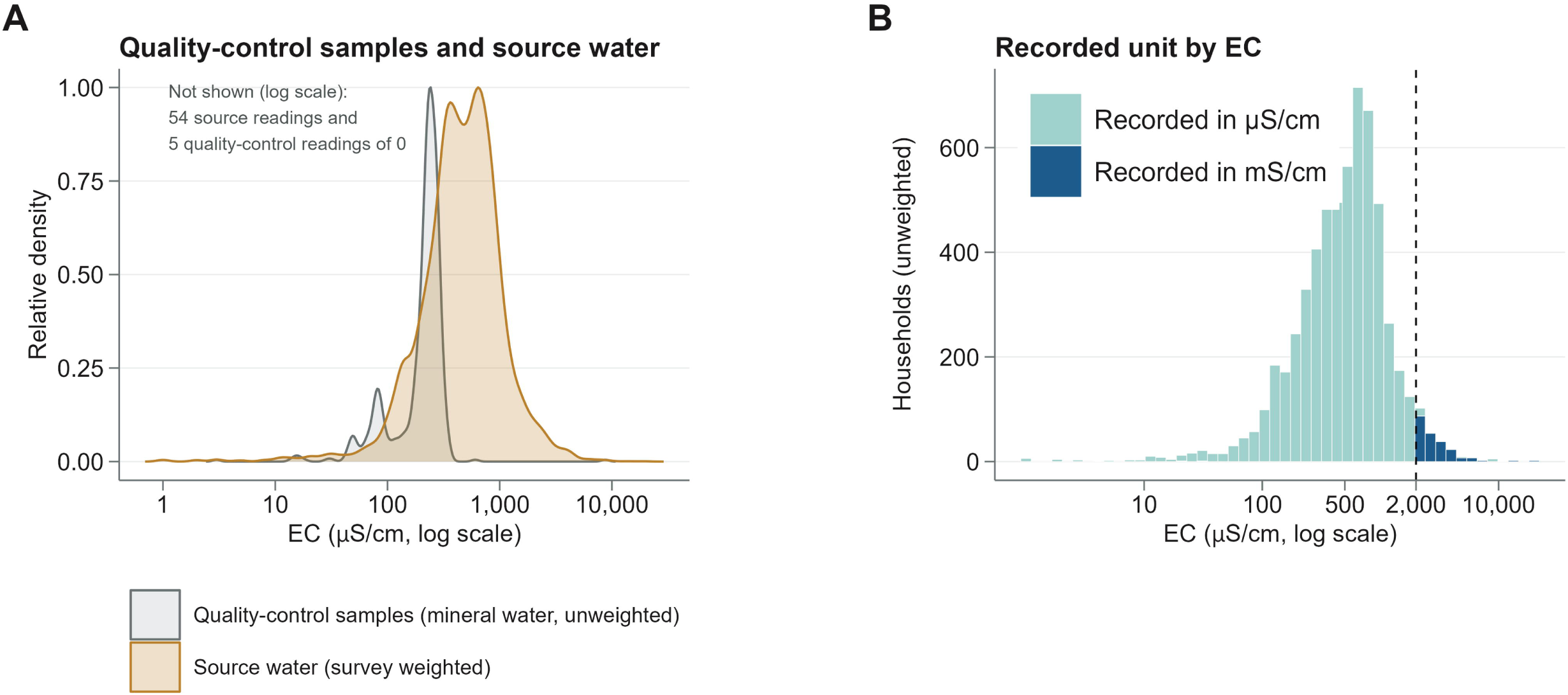
Measurement quality. (A) Density of log10 EC in source water (survey weighted) and in 605 quality-control samples of bottled mineral water (unweighted). Readings of 0 cannot be shown on the log scale: 54 source readings and 5 quality-control readings. (B) Source readings by recorded unit, unweighted counts. Of readings of 2,000 µS/cm or more, 95% were recorded in mS/cm, consistent with an auto-ranging meter. Bangladesh MICS 2025. (TIF)

**S3 Fig.**
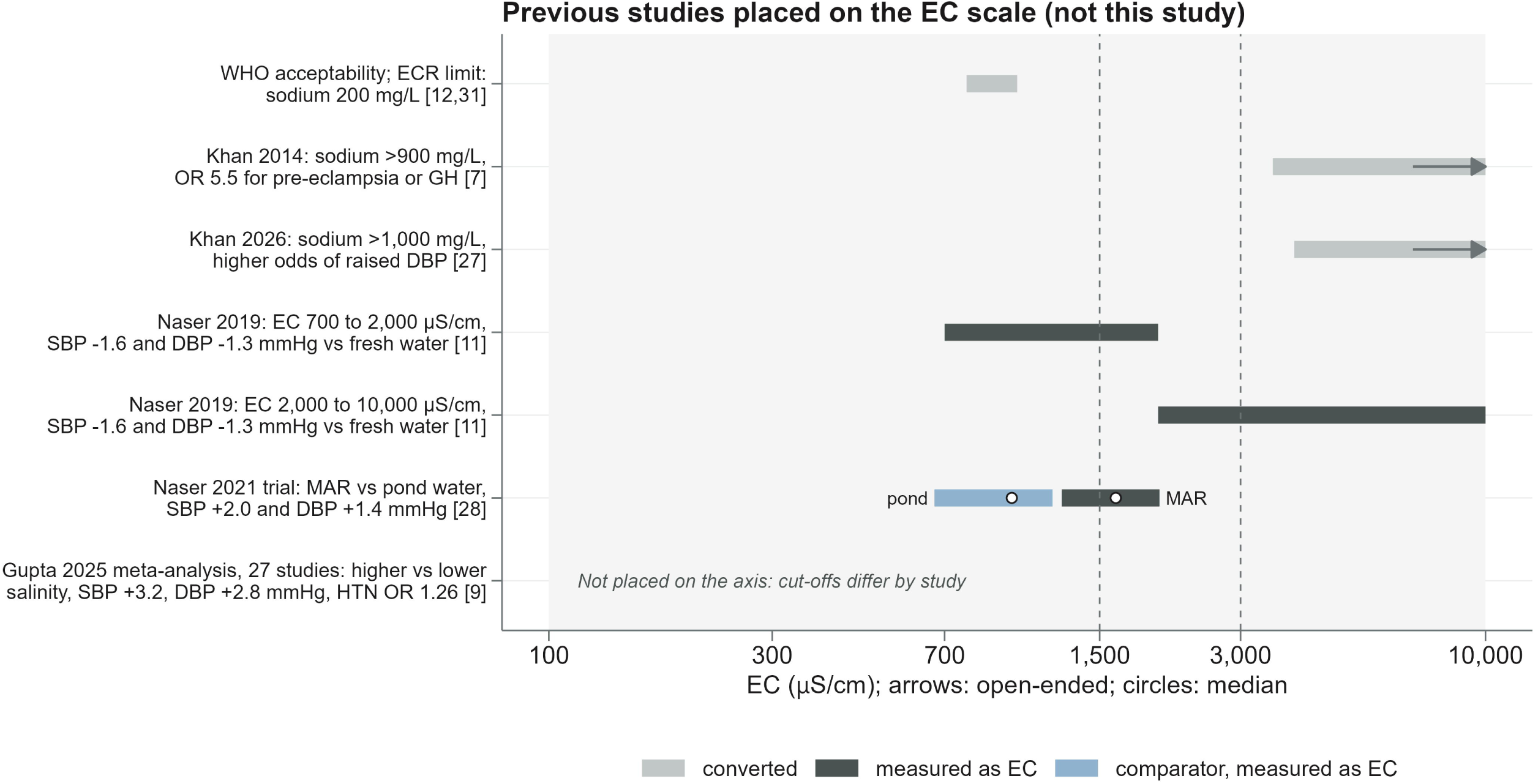
Previous health studies placed on the EC scale (not results of this study). Ranges are converted from sodium (light) or measured as EC (dark); arrows mark open-ended categories. Dashed lines: the survey classes at 1,500 and 3,000 µS/cm. Sodium was converted from EC as 0.4 x 0.64 x EC, or as 0.20 x EC from the ratio of mean sodium to mean EC in Dacope [27]. Both apply only to coastal sodium-chloride water and are indicative only. Naser 2019 (main text reference 11): SBP-1.55 and-1.58 mmHg and DBP-1.26 and-1.28 mmHg for EC 700 to 2,000 and 2,000 to 10,000 µS/cm; both round to-1.6 and-1.3. Naser 2021 (reference 28): bars, interquartile range of EC in recharge and pond water; circles, medians. Gupta 2025 (reference 9) is not on the axis. GH, gestational hypertension; HTN, hypertension; MAR, managed aquifer recharge; OR, odds ratio; SBP, DBP, systolic and diastolic blood pressure. Numbers in brackets in the figure are main text reference numbers. (TIF)

## Notes

### Competing Interest Statement

The authors have declared no competing interest.

### Author Declarations

This study is a secondary analysis of de-identified data from the Bangladesh Multiple Indicator Cluster Survey (MICS) 2025. Ethics committee/IRB of International Centre for Diarrhoeal Disease Research, Bangladesh (icddr,b) gave ethical approval for the MICS 2025 survey data collection (ref: PR-24113). No separate ethics approval was obtained for this secondary analysis.

### Summary of Updates

Added a regional analysis of deep against shallow tube wells. The difference varied by region, and the abstract and conclusions now say so. Added the meter calibration limitation and the sensitivity range (4.5% to 6.2% in most scenarios) to the abstract. Identified six discordant readings as probable recording errors, with a sensitivity check. Simplified the arsenic analysis to a cross-classification of high EC and arsenic. Clarified zone totals, source type labels and the wording of several findings. Updated Supporting Information (S1 to S6 Tables) to match.

## References

1. Mueller W, Zamrsky D, Essink GO, Fleming LE, Deshpande A, Makris KC, et al. Saltwater intrusion and human health risks for coastal populations under 2050 climate scenarios. Sci Rep. 2024;14(1):15881. doi: 10.1038/s41598-024-66956-4

2. Khan AE, Ireson A, Kovats S, Mojumder SK, Khusru A, Rahman A, et al. Drinking water salinity and maternal health in coastal Bangladesh: implications of climate change. Environ Health Perspect. 2011;119(9):1328–32. doi: 10.1289/ehp.1002804

3. Tsai C, Hoque MA, Vineis P, Ahmed KM, Butler AP. Salinisation of drinking water ponds and groundwater in coastal Bangladesh linked to tropical cyclones. Sci Rep. 2024;14(1):5211. doi: 10.1038/s41598-024-54446-6

4. Nahian MA, Ahmed A, Lázár AN, Hutton CW, Salehin M, Streatfield PK. Drinking water salinity associated health crisis in coastal Bangladesh. Elementa. 2018;6:2. doi: 10.1525/elementa.143

5. Costopoulos E, Imamura A, Khan N, Butler A, Millett C, Hoque MA, et al. Adverse health outcomes associated with drinking highly saline water: a systematic review. Eur J Epidemiol. 2025;40(11):1307–22. doi: 10.1007/s10654-025-01307-9

6. Talukder MRR, Rutherford S, Phung D, Islam MZ, Chu C. The effect of drinking water salinity on blood pressure in young adults of coastal Bangladesh. Environ Pollut. 2016;214:248–54. doi: 10.1016/j.envpol.2016.03.074

7. Khan AE, Scheelbeek PFD, Shilpi AB, Chan Q, Mojumder SK, Rahman A, et al. Salinity in drinking water and the risk of (pre)eclampsia and gestational hypertension in coastal Bangladesh: a case-control study. PLoS One. 2014;9(9):e108715. doi: 10.1371/journal.pone.0108715

8. Scheelbeek PFD, Chowdhury MAH, Haines A, Alam DS, Hoque MA, Butler AP, et al. Drinking water salinity and raised blood pressure: evidence from a cohort study in coastal Bangladesh. Environ Health Perspect. 2017;125(5):057007. doi: 10.1289/EHP659

9. Gupta RD, Kunutsor SK, Lopez-Salamanca DE, Mercy FT, Nawal N, Tejada CE, et al. Association of drinking water salinity with elevated blood pressure and risk of hypertension among coastal and other populations: a systematic review and meta-analysis of observational studies. BMJ Glob Health. 2025;10(12):e018543. doi: 10.1136/bmjgh-2024-018543

10. Talukder MRR, Rutherford S, Huang C, Phung D, Islam MZ, Chu C. Drinking water salinity and risk of hypertension: a systematic review and meta-analysis. Arch Environ Occup Health. 2017;72(3):126–38. doi: 10.1080/19338244.2016.1175413

11. Naser AM, Rahman M, Unicomb L, Doza S, Gazi MS, Alam GR, et al. Drinking water salinity, urinary macro-mineral excretions, and blood pressure in the southwest coastal population of Bangladesh. J Am Heart Assoc. 2019;8(9):e012007. doi: 10.1161/JAHA.119.012007

12. World Health Organization. Guidelines for drinking-water quality. 4th ed, incorporating the first and second addenda. Geneva: World Health Organization; 2022.

13. Bangladesh Bureau of Statistics, UNICEF Bangladesh. Bangladesh National Drinking Water Quality Survey of 2009. Dhaka: Bangladesh Bureau of Statistics; 2011.

14. Ayers JC, Goodbred S, George G, Fry D, Benneyworth L, Hornberger G, et al. Sources of salinity and arsenic in groundwater in southwest Bangladesh. Geochem Trans. 2016;17(1):4. doi: 10.1186/s12932-016-0036-6

15. Rakib MA, Sasaki J, Matsuda H, Quraishi SB, Mahmud MJ, Bodrud-Doza M, et al. Groundwater salinization and associated co-contamination risk increase severe drinking water vulnerabilities in the southwestern coast of Bangladesh. Chemosphere. 2020;246:125646. doi: 10.1016/j.chemosphere.2019.125646

16. Akter S. Impact of drinking water salinity on children’s education: empirical evidence from coastal Bangladesh. Sci Total Environ. 2019;690:1331–41. doi: 10.1016/j.scitotenv.2019.06.458

17. Bain R, Johnston R, Khan S, Hancioglu A, Slaymaker T. Monitoring drinking water quality in nationally representative household surveys in low-and middle-income countries: cross-sectional analysis of 27 Multiple Indicator Cluster Surveys 2014 to 2020. Environ Health Perspect. 2021;129(9):097010. doi: 10.1289/EHP8459

18. Bangladesh Bureau of Statistics, UNICEF Bangladesh. Progotir Pathey, Bangladesh Multiple Indicator Cluster Survey 2025, survey findings report. Dhaka: Bangladesh Bureau of Statistics; 2026.

19. Sarker MMR, Van Camp M, Hossain D, Islam M, Ahmed N, Karim MM, et al. Groundwater salinization and freshening processes in coastal aquifers from southwest Bangladesh. Sci Total Environ. 2021;779:146339. doi: 10.1016/j.scitotenv.2021.146339

20. Ministry of Water Resources, Government of the People’s Republic of Bangladesh. Coastal Zone Policy 2005. Dhaka: Ministry of Water Resources; 2005.

21. Runfola D, Anderson A, Baier H, Crittenden M, Dowker E, Fuhrig S, et al. geoBoundaries: a global database of political administrative boundaries. PLoS One. 2020;15(4):e0231866. doi: 10.1371/journal.pone.0231866

22. Bangladesh Bureau of Statistics, Bangladesh Institute of Development Studies. Population and Housing Census 2022: post enumeration check adjusted population. Dhaka: Bangladesh Bureau of Statistics; 2023.

23. Bangladesh Bureau of Statistics. Population and Housing Census 2022, national report (volume I). Dhaka: Bangladesh Bureau of Statistics; 2023.

24. Shuvo SD, Zahid MA, Rahman MM, Parvin R. Exploring the impact of soil and water salinity on dietary behavior and health risk of coastal communities in Bangladesh. J Water Health. 2020;18(6):1110–23. doi: 10.2166/wh.2020.145

25. Ravenscroft P, McArthur JM, Hoque MA. Stable groundwater quality in deep aquifers of Southern Bangladesh: the case against sustainable abstraction. Sci Total Environ. 2013;454–455:627-38. doi: 10.1016/j.scitotenv.2013.02.071

26. Scheelbeek PFD, Khan AE, Mojumder S, Elliott P, Vineis P. Drinking water sodium and elevated blood pressure of healthy pregnant women in salinity-affected coastal areas. Hypertension. 2016;68(2):464–70. doi: 10.1161/HYPERTENSIONAHA.116.07743

27. Khan AE, Costopoulos E, Mojumder SK, Shilpi AB, Hoque MA, Butler A, et al. Drinking water salinity and blood pressure in coastal Bangladesh: follow-up of a population-based cohort in a climate-vulnerable region. BMJ Open. 2026;16(7):e116453. doi: 10.1136/bmjopen-2026-116453

28. Naser AM, Doza S, Rahman M, Unicomb L, Ahmed KM, Anand S, et al. Consequences of access to water from managed aquifer recharge systems for blood pressure and proteinuria in south-west coastal Bangladesh: a stepped-wedge cluster-randomized trial. Int J Epidemiol. 2021;50(3):916–28. doi: 10.1093/ije/dyaa098

29. Naser AM, Rahman M, Unicomb L, Parvez SM, Islam S, Doza S, et al. Associations of drinking rainwater with macro-mineral intake and cardiometabolic health: a pooled cohort analysis in Bangladesh, 2016 to 2019. NPJ Clean Water. 2020;3(1):20. doi: 10.1038/s41545-020-0067-5

30. Ministry of Environment and Forests, Government of the People’s Republic of Bangladesh. The Environment Conservation Rules, 1997. Schedule 3(B): standards for drinking water. Dhaka: Ministry of Environment and Forests; 1997.

31. Ministry of Environment, Forest and Climate Change, Government of the People’s Republic of Bangladesh. Environment Conservation Rules, 2023 (S.R.O. No. 53-Law/2023). Schedule 2(B): standards for drinking water. Bangladesh Gazette, Extraordinary. 2023 Mar 5:3063-5.

32. Biswas S, Chattopadhyay A, Schilling K, Das A. Investigating the association between groundwater contaminants and hypertension risk in India: a machine learning-based analysis. J Expo Sci Environ Epidemiol. 2026;36(1):125–42. doi: 10.1038/s41370-025-00776-0

33. Sarkar S, Das K, Mukherjee A. Groundwater salinity across India: predicting occurrences and controls by field-observations and machine learning modeling. Environ Sci Technol. 2024;58(8):3953–65. doi: 10.1021/acs.est.3c06525

34. Le H, Key K, Steckler MS, Sazeed N, Person M, Bhuiyan A, et al. Buried deep freshwater reserves beneath salinity-stressed coastal Bangladesh. Nat Commun. 2025;16(1):10740. doi: 10.1038/s41467-025-65770-4

