## Supplementary material for "Source-water salinity measured by electrical conductivity in Bangladesh, 2025: a nationally representative cross-sectional survey": S1_Checklist.docx

### S1 Checklist. STROBE checklist for cross-sectional studies

Manuscript: “Source-water salinity measured by electrical conductivity in Bangladesh, 2025: a nationally representative cross-sectional survey”. Location gives the section of the manuscript or the supporting information item. Status: **Met**, **Partly** or **Not applicable**.

| Item | Recommendation | Location in the manuscript | Status |
| --- | --- | --- | --- |
| 1a | Study design in the title or abstract | Title (“cross-sectional survey”); Abstract, Methods | Met |
| 1b | Informative and balanced abstract | Abstract (Background, Objectives, Methods, Results, Conclusions; 464 words) | Met |
| 2 | Scientific background and rationale | Introduction, paragraphs 1 to 5 (salinisation, health evidence, gap, MICS 2025) | Met |
| 3 | Objectives | Introduction, last paragraph (one overall objective, then specific aims; month exploratory) | Met |
| 4 | Key elements of study design early in the paper | Methods, Data, sampling and measurement, paragraph 1 (secondary analysis of a stratified two-stage cluster survey) | Met |
| 5 | Setting, locations and relevant dates | Methods, Data, sampling and measurement (all 64 districts; fieldwork 24 February to 30 June 2025) | Met |
| 6a | Eligibility criteria, sources and methods of selection | Methods, Data, sampling and measurement (2 of 20 households per cluster chosen at random for water testing); S1 Table | Met |
| 7 | Outcomes, exposures, predictors, confounders and effect modifiers | Methods, Variables (outcomes, three measures, household characteristics, place, source and arsenic); Statistical analysis, Source type (adjustment set); S2 Table | Met |
| 8 | Data sources and measurement, and comparability | Methods, Data, sampling and measurement (measurement procedure, units, field teams and training); Variables; Sensitivity and exploratory analyses (quality control, low readings) | Met |
| 9 | Efforts to address potential sources of bias | Methods, Sensitivity and exploratory analyses; S1 Table (non-measurement model); S3 Table (measurement and missing-data scenarios); S6 Table | Met |
| 10 | How the study size was arrived at | Methods, Data, sampling and measurement (sample size set by the survey; precision reported instead of power); Results, first paragraph (6,298 selected, 6,111 interviewed, 6,040 with EC) | Met |
| 11 | Handling of quantitative variables | Methods, Variables (cut-offs at 700, 1,000, 1,500, 2,000 and 3,000 µS/cm); Statistical analysis (log10 EC, geometric mean ratios); S2 Table (wealth coding, elevation and rainfall terms) | Met |
| 12a | Statistical methods, including control for confounding | Methods, Statistical analysis (prevalence ratios, linear models on log10 EC, adjustment sets and rationale); S2 Table | Met |
| 12b | Subgroups and interactions | Methods, Statistical analysis (coastal zone and other districts analysed separately; well type by coastal region interaction; division and district estimates); S2 and S6 Tables | Met |
| 12c | Missing data | Methods, Sensitivity and exploratory analyses (bounds for missing readings; complete cases otherwise); Table 1, Missing column; S3 Table | Met |
| 12d | Analytical methods taking account of sampling strategy | Methods, Data (water-quality weight, strata, clusters); Statistical analysis (Taylor linearisation) | Met |
| 12e | Sensitivity analyses | Methods, Sensitivity and exploratory analyses; S3 and S6 Tables | Met |
| 13a | Numbers of individuals at each stage | Results, first paragraph; S1 Table | Met |
| 13b | Reasons for non-participation | S1 Table note (no recorded result 65, refused 4, no water 2) | Met |
| 13c | Flow diagram | S1 Table gives the flow as a table | Partly: no diagram; a table is acceptable for a survey subsample |
| 14a | Characteristics of participants, exposures and confounders | Results, first paragraph; Table 1 | Met |
| 14b | Number of participants with missing data for each variable | Table 1, Missing (n) column | Met |
| 15 | Numbers of outcome events or summary measures | Table 2; Results, paragraph on prevalence and population burden; Fig 1 | Met |
| 16a | Unadjusted and adjusted estimates with precision, and confounders adjusted for | Results (prevalence ratio 2.85 crude, 0.79 district adjusted; GMR 1.09 crude, 0.82 primary); Methods (adjustment set); S6 Table | Met |
| 16b | Category boundaries when continuous variables are categorised | Table 2 (cut-offs); S5 Table, Panel G (EC bands) | Met |
| 16c | Translation of relative risk into absolute risk | Table 2 and Results (people above each cut-off, in millions) | Met |
| 17 | Other analyses (subgroups, interactions, sensitivity) | Results (measurement scenarios, deep and shallow tube wells with the regional interaction, exploratory month); S3 and S6 Tables | Met |
| 18 | Key results with reference to study objectives | Discussion, paragraphs 1 and 2 | Met |
| 19 | Limitations, with direction and magnitude of bias | Discussion, Strengths and limitations (five limitations; direction given for missing readings, counts, season and well-depth misclassification) | Met |
| 20 | Cautious overall interpretation | Discussion; Implications (stated as not tested); Conclusions | Met |
| 21 | Generalisability (external validity) | Discussion, Strengths and limitations (last sentence: tested sources of Bangladeshi households, dry and pre-monsoon season of 2025) | Met |
| 22 | Funding and role of funders | Entered in the submission form (no specific funding) | Met |
