## Supplementary material for "Source-water salinity measured by electrical conductivity in Bangladesh, 2025: a nationally representative cross-sectional survey": S1_Table.docx

### S1 Table. Sample flow, non-measurement and analysis samples

**Panel A. Households selected for source testing, by division**

| Division | Selected | Completed interview | With EC reading | % of completed | Not measured | Adjusted odds ratio of non-measurement (95% CI) |
| --- | --- | --- | --- | --- | --- | --- |
| Barishal | 564 | 540 | 535 | 99.1 | 5 | 1.95 (0.61 to 6.26) |
| Chattogram | 1,128 | 1,084 | 1,078 | 99.4 | 6 | 1.07 (0.36 to 3.20) |
| Dhaka | 1,410 | 1,373 | 1,366 | 99.5 | 7 | 1 (reference) |
| Khulna | 940 | 912 | 872 | 95.6 | 40 | 10.62 (4.66 to 24.21) |
| Mymensingh | 376 | 363 | 358 | 98.6 | 5 | 2.97 (0.92 to 9.59) |
| Rajshahi | 752 | 738 | 735 | 99.6 | 3 | 0.93 (0.24 to 3.65) |
| Rangpur | 752 | 735 | 733 | 99.7 | 2 | 0.59 (0.12 to 2.88) |
| Sylhet | 376 | 366 | 363 | 99.2 | 3 | 1.64 (0.42 to 6.45) |
| Total | 6,298 | 6,111 | 6,040 | 98.8 | 71 |  |

*Logistic model of non-measurement among the 6,111 households with a completed interview, mutually adjusted for division, residence, wealth quintile and household size. Other odds ratios: urban 1.33 (0.77 to 2.29) and household size 1.05 per member (0.91 to 1.20); all wealth quintile intervals included 1. Likelihood ratio tests: division P = 4 × 10^-14^; residence, wealth and household size together P = 0.46 without division and P = 0.08 with division. Of the 71 not measured, 65 had no recorded source-test result, 4 refused permission and 2 had no water at the source. The report’s 6,010 completed source tests are households with a source E. coli sample; 30 further households had an EC reading without one and are analysed.*

**Panel B. Unit and sample size of each analysis**

| Analysis | Unit | n |
| --- | --- | --- |
| Prevalence, population burden, source type, wealth, arsenic cross-classification | Household (members for population estimates) | 6,040 (source “other” 167) |
| Month and depth log models | Household, zero readings excluded | 5,986; coastal 1,746; coastal tube wells 1,433 |
| Variance components | Household, 41 measurers | 5,986 |
| Moran’s I, spatial surface | Cluster (mean of one or two tests) | 3,124 |
| How households obtained water | Household, coastal zone | 1,762 with a known source location; 1,765 for treatment; 1,769 for wet-season source |
| Children under 5, pregnant women | Household | 2,058 with a child under 5; 284 with a currently pregnant woman; 897 with no interviewed woman aged 15 to 49 |
| Measurement quality | Test | 605 quality-control samples; 54 zero readings |
