## Supplementary material for "Source-water salinity measured by electrical conductivity in Bangladesh, 2025: a nationally representative cross-sectional survey": S2_Table.docx

### S2 Table. Model specifications

Rows follow the study aims (prevalence, geographic patterns, source type, co-occurrence with arsenic), then sensitivity and exploratory analyses. All models use the water-quality weight for the source test, strata and clusters (Taylor linearisation) unless marked unweighted. Wealth quintile is categorical unless stated. Elevation and rainfall are cluster-level and linear. FE, fixed effects.

| Title theme | Analysis | Outcome | Exposure (reference) | Adjustment | Sample (n) | Model |
| --- | --- | --- | --- | --- | --- | --- |
| Prevalence | Prevalence and population counts | EC above each cut-off | None | None | 6,040 households | Survey proportion, logit interval; members weighted by household size |
| Prevalence | Factors associated with high EC (urban ratio in S5 Table) | EC above 700 or 1,500 µS/cm | Residence (rural), wealth (poorest), tested source (shallow tube well), division (Dhaka) | Mutually adjusted | 6,040; Sylhet excluded at 1,500 | Quasi-Poisson, log link |
| Prevalence | Wealth trend | EC above 1,500 µS/cm | Wealth quintile, linear, per quintile | Residence, tested source, division | National without Sylhet; coastal zone | Quasi-Poisson, log link |
| Prevalence | How households obtained water | Main source not in dwelling or yard | EC above 1,500 µS/cm (at or below) | Residence, wealth and division; or residence, wealth and district | 1,762 coastal households with known location | Quasi-Poisson, log link |
| Geographic patterns | Coastal against other districts | EC above 700, 1,500 or 3,000 µS/cm | Coastal zone (other districts) | Crude; residence and wealth | 6,040 | Quasi-Poisson, log link |
| Geographic patterns | Spatial structure | Cluster mean of log10 EC | None | Residence, wealth, elevation, rainfall, division; thin plate spline (k = 60) | 3,124 clusters | Gaussian additive model (REML), unweighted; Moran’s I of residuals |
| Source type | Deep against shallow, prevalence | EC above 1,500 µS/cm | Deep tube well (shallow) | Crude; division; district | 4,894 tube wells | Quasi-Poisson, log link |
| Source type | Deep against shallow, prevalence difference | EC above 1,500 µS/cm | Deep tube well (shallow) | None | 1,436 coastal tube wells | Linear, identity link |
| Source type | **Deep against shallow, primary** | log10 EC | Deep tube well (shallow) | District FE, elevation, rainfall, residence, wealth | EC above 0: coastal 1,433; other districts 3,435 | Linear on log10 EC; GMR = 10^coefficient |
| Source type | Deep against shallow, by coastal region (S6 Table) | log10 EC | Deep tube well (shallow), within each region | Primary covariates plus a well type by region interaction | Coastal tube wells: Khulna and Barishal 826; Chattogram 432; Gopalganj and Shariatpur 175 | Linear on log10 EC; design-based Wald test of the interaction |
| Source type | Deep against shallow, sensitivity (S6 Table) | log10 EC | Deep tube well (shallow) | Crude; residence, wealth and division or district; primary plus measurer FE; upazila FE in place of district; splines (3 df) for elevation and rainfall | As primary; upazila model identified from 851 coastal households | Linear on log10 EC |
| Co-occurrence with arsenic | Cross-classification | EC above 1,500 µS/cm and arsenic above 10 µg/L | None | None | 6,040 | Survey proportions; members weighted by household size |
| Sensitivity (low readings) | Pattern of readings of 20 µS/cm or less | Reading of 20 µS/cm or less | Month, division, zone, tested source; district; measurer | None | 6,040 | Rao-Scott F test, survey weighted; Monte Carlo chi-square, unweighted, for district and measurer |
| Sensitivity (discordant readings) | Six readings in Rangpur division excluded | EC above 1,500 and 3,000 µS/cm | None | None | 6,034 | Survey proportion, logit interval |
| Exploratory (month) | **Month, primary (exploratory)** | log10 EC | April; May to June (February to March) | Primary depth covariates plus tested source | EC above 0, July excluded: coastal 1,746; other districts 4,238 | Linear on log10 EC |
| Exploratory (month) | Month within source type | log10 EC | As above | Primary depth covariates | Coastal deep 912, shallow 521, piped 221 | Linear on log10 EC |
| Sensitivity (measurer) | Variance components | log10 EC | None | Random intercepts for district (or upazila), measurer and cluster | 5,986 | Linear mixed model, unweighted |

*Zero readings (54) were excluded from all log models; readings of 20 µS/cm or less were also excluded in sensitivity analyses (S6 Table, Panel C).*
