## Supplementary material for "Source-water salinity measured by electrical conductivity in Bangladesh, 2025: a nationally representative cross-sectional survey": S3_Table.docx

### S3 Table. Sensitivity of the main estimates to measurement, missing data and measurer

| Check | Result |
| --- | --- |
| **A. Measurement** |  |
| Distribution of EC readings (6,040 source readings) | Range 0 to 20,010 µS/cm. Percentiles 1st, 5th, 10th, 25th, 50th, 75th, 90th, 95th and 99th: 1, 105, 153, 285, 462, 742, 1,090, 1,527 and 3,150 µS/cm. 54 readings were zero, 52 were 1 to 20 µS/cm, 432 exceeded 1,500 and 82 exceeded 3,000 µS/cm |
| Shift all readings by -100 / +100 µS/cm | Above 700: 21.2% / 37.2%. Above 1,500: 4.5% / 6.0%. Above 3,000: 1.01% / 1.17% |
| Multiply all readings by 0.8 / 0.9 / 1.1 / 1.2 | Above 1,500: 3.4% / 4.2% / 6.3% / 7.3%. Above 3,000: 0.66% / 0.90% / 1.37% / 1.66% |
| Readings of 20 µS/cm or less | 106 readings: 54 zero (from 46 clusters; 53 recorded in µS/cm) and 52 of 1 to 20 µS/cm. By tested source: shallow tube wells 48, deep tube wells 14, piped 29, other 15 (9.0% of that group). Of 97 with a cluster-mate, the mate had a median EC of 403 µS/cm and 20 mates also read 20 or less. 11 of 605 quality-control samples also read below 20 µS/cm, 5 of them zero |
| All readings of 20 µS/cm or less set to missing, or assumed above each cut-off | As measured: above 700 28.14%, above 1,500 5.29%, above 3,000 1.05%. Set to missing: 28.66%, 5.39%, 1.07%. Assumed above the cut-off: above 1,500 7.08%, above 3,000 2.84% |
| Pattern of readings of 20 µS/cm or less | By month of testing, February to June: 1.3%, 2.0%, 1.8%, 1.9%, 0.8% of tests (survey-weighted P = 0.35). By division: 0.9% to 2.3% (P = 0.14). Coastal zone 2.7% against 1.4% elsewhere (P < 0.001). 40 of 64 districts had at least one (unweighted Monte Carlo P < 0.001). 28 of 41 measurers recorded at least one; five measurers, with 12.1% of tests, recorded 48.1% (P < 0.001) |
| Six readings in Rangpur division (five in Panchagarh, one in Gaibandha) of 3,338 to 9,906 µS/cm, each at least 9 times the reading of the other household in its cluster | Above 3,000 µS/cm: 1.00% without them, 1.05% with them. Above 1,500 µS/cm: 5.25% against 5.29% |
| Temperature: uncompensated-meter scenario, water at ambient / half the departure | National share above 700: 25.5% / 26.8%; above 1,500: 4.7% / 4.94%; above 3,000: 0.95% / 1.00% (as measured 28.1%, 5.29%, 1.05%). Coastal month ratio (primary): 1.10 (0.92 to 1.31) / 1.13 (0.95 to 1.35) |
| **B. Missing data and tested source** |  |
| 71 unmeasured households in completed interviews, all below / all above 1,500 µS/cm | Above 1,500: 5.20% / 6.23% (observed 5.29%); Khulna 13.8% / 18.2% (observed 14.5%). Above 3,000: 1.03% / 2.06% (observed 1.05%) |
| All 258 selected households without a reading, including 187 without a completed interview | 5.08% / 9.17% |
| Tested source was also the main source (5,175 households; 85.5% weighted) | Above 1,500: 5.26% (4.67 to 5.93) of households and 5.31% (4.68 to 6.02) of members. Applied to all 169.8 million people this gives 9.0 million (8.0 to 10.2); within the matching-source households alone (85.2% of members) it gives 7.7 million. Above 3,000: 1.06% (0.78 to 1.44) of households; 1.8 million applied to all people, about 1.5 million within matching-source households |
| **C. Measurer** |  |
| Measurers; households per measurer, median (range) | 41; 149 (121 to 158); 9 worked in more than one division |
| Variance of log10 EC: district, measurer, cluster, residual (unweighted mixed model) | 27%, 18%, 18%, 37% |
| **D. Replication in Stata 17** |  |
| Primary estimates | Above 1,500: 5.29% (4.69 to 5.90). Above 3,000: 1.05% (0.75 to 1.35). Coastal deep against shallow, primary specification: GMR 0.819 |

*Percentages are household weighted. Missing-data bounds apply each division’s unmeasured share of selected households to the supplied weights, which already adjust for nonresponse. The additive and multiplicative error scenarios are chosen hypothetical scenarios, not validated bounds. Stata intervals for proportions are Wald intervals, so they differ slightly from the logit intervals in the main text. The temperature scenarios assume an uncompensated meter; the actual compensation is not documented. The six discordant readings were identified by inspection in Rangpur division, where EC is otherwise low. Pairs with a similar ratio in the coastal zone were not excluded, because high readings are plausible there.*
