## Supplementary material for "Source-water salinity measured by electrical conductivity in Bangladesh, 2025: a nationally representative cross-sectional survey": S4_Table.docx

### S4 Table. Estimates by division, zone and district

**Panel A. Divisions and zones**

| Group | Households (n) | Households above 1,500, % (95% CI) | Members above 1,500, % (95% CI) | Households above 3,000, % (95% CI) | People above 1,500, thousands (95% CI) | Median EC (IQR) |
| --- | --- | --- | --- | --- | --- | --- |
| Barishal | 535 | 15.0 (12.0 to 18.6) | 13.6 (10.8 to 17.1) | 3.0 (1.9 to 4.7) | 1,270 (1,000 to 1,590) | 855 (648 to 1,165) |
| Khulna | 872 | 14.5 (12.1 to 17.2) | 14.5 (11.9 to 17.6) | 3.6 (2.3 to 5.5) | 2,590 (2,120 to 3,130) | 755 (590 to 1,015) |
| Chattogram | 1,078 | 7.7 (5.8 to 10.2) | 7.6 (5.7 to 10.2) | 1.2 (0.5 to 2.7) | 2,610 (1,940 to 3,480) | 439 (258 to 757) |
| Dhaka | 1,366 | 3.7 (3.0 to 4.7) | 3.9 (3.1 to 4.8) | 0.5 (0.2 to 0.9) | 1,770 (1,420 to 2,200) | 461 (318 to 730) |
| Mymensingh | 358 | 1.7 (0.6 to 4.5) | 1.6 (0.6 to 4.5) | 1.0 (0.2 to 4.1) | 208 (74 to 575) | 334 (263 to 462) |
| Rajshahi | 735 | 0.9 (0.4 to 2.1) | 1.0 (0.4 to 2.4) | 0 (0 to 0.5) | 206 (86 to 490) | 541 (377 to 694) |
| Rangpur | 733 | 1.2 (0.6 to 2.2) | 1.3 (0.7 to 2.5) | 0.4 (0.2 to 1.0) | 232 (120 to 446) | 251 (151 to 392) |
| Sylhet | 363 | 0 (0 to 1.0) | 0 (0 to 1.0) | 0 (0 to 1.0) | 0 | 219 (138 to 352) |
| 19 coastal-zone districts | 1,771 | 14.1 (12.4 to 16.1) | 13.8 (12.0 to 15.8) | 3.1 (2.2 to 4.2) | 6,210 (5,390 to 7,130) | 684 (400 to 1,065) |
| Other 45 districts | 4,269 | 2.1 (1.7 to 2.7) | 2.1 (1.6 to 2.6) | 0.3 (0.2 to 0.6) | 2,580 (2,020 to 3,280) | 410 (271 to 643) |
| Bangladesh | 6,040 | 5.3 (4.7 to 5.9) | 5.3 (4.7 to 6.0) | 1.05 (0.79 to 1.40) | 8,980 (7,940 to 10,100) | 462 (285 to 742) |

*Household percentages use the water-quality weight; member percentages that weight times household size. People = member-weighted proportion x subgroup reference population, rounded to three significant figures. Reference populations are the 2022 post enumeration check counts for divisions, 45.0 million for the coastal zone and 124.8 million for the other districts. Subgroup counts use different reference populations and do not sum exactly to the national count. “0 (0 to x)”: no sampled household exceeded the cut-off; x is the exact binomial upper limit, ignoring clustering. Member-weighted shares above 1,500 µS/cm by division reproduce Table WS.1.6S1 of the survey report (main text ref 18). Median EC in µS/cm, household weighted.*

**Panel B. Districts**

| District | Coastal zone | Households (n) | Above 700, % (95% CI) | Above 1,500, % (95% CI) | Above 3,000, % (95% CI) | Design effect at 1,500 |
| --- | --- | --- | --- | --- | --- | --- |
| Bagerhat | Yes | 80 | 62.5 (47.7 to 75.3) | 55.0 (41.3 to 67.9) | 10.9 (5.5 to 20.5) | 1.5 |
| Bandarban | No | 86 | 2.2 (0.5 to 8.8) | 1.1 (0.1 to 8.2) | 0 (0 to 4.2) | 1.0 |
| Barguna | Yes | 92 | 95.2 (87.8 to 98.2) | 30.4 (19.3 to 44.4) | 0 (0 to 3.9) | 1.7 |
| Barishal | Yes | 88 | 77.8 (64.9 to 86.9) | 4.3 (1.3 to 13.2) | 0 (0 to 4.1) | 1.3 |
| Bhola | Yes | 88 | 19.0 (11.0 to 30.6) | 0 (0 to 4.1) | 0 (0 to 4.1) | n/a |
| Bogura | No | 92 | 19.8 (11.3 to 32.3) | 1.2 (0.2 to 8.4) | 0 (0 to 3.9) | 1.1 |
| Brahmanbaria | No | 87 | 23.4 (13.2 to 38.2) | 7.7 (2.7 to 20.0) | 0 (0 to 4.2) | 1.8 |
| Chandpur | Yes | 92 | 37.5 (26.3 to 50.1) | 5.3 (1.8 to 14.4) | 0 (0 to 3.9) | 1.4 |
| Chapai Nawabganj | No | 90 | 29.2 (19.1 to 41.9) | 1.1 (0.1 to 8.0) | 0 (0 to 4.0) | 1.0 |
| Chattogram | Yes | 180 | 20.2 (13.9 to 28.4) | 2.5 (0.8 to 7.6) | 0 (0 to 2.0) | 1.5 |
| Chuadanga | No | 89 | 56.5 (44.4 to 67.9) | 4.1 (1.5 to 10.8) | 0.9 (0.1 to 6.8) | 0.9 |
| Cox’s Bazar | Yes | 89 | 16.0 (9.1 to 26.7) | 3.2 (1.0 to 10.1) | 0 (0 to 4.1) | 1.0 |
| Cumilla | No | 90 | 27.0 (17.9 to 38.7) | 8.0 (3.4 to 17.7) | 1.2 (0.2 to 8.6) | 1.3 |
| Dhaka | No | 280 | 13.1 (8.5 to 19.7) | 0.7 (0.2 to 2.8) | 0.3 (0.0 to 2.1) | 1.0 |
| Dinajpur | No | 93 | 5.9 (2.2 to 14.8) | 0 (0 to 3.9) | 0 (0 to 3.9) | n/a |
| Faridpur | No | 91 | 72.3 (59.3 to 82.4) | 10.5 (5.4 to 19.5) | 0.9 (0.1 to 6.4) | 1.1 |
| Feni | Yes | 92 | 29.7 (19.1 to 43.1) | 11.1 (5.1 to 22.5) | 1.3 (0.2 to 9.1) | 1.6 |
| Gaibandha | No | 91 | 14.1 (8.0 to 23.7) | 2.5 (0.8 to 7.9) | 0.8 (0.1 to 5.9) | 0.8 |
| Gazipur | No | 92 | 0 (0 to 3.9) | 0 (0 to 3.9) | 0 (0 to 3.9) | n/a |
| Gopalganj | Yes | 90 | 86.9 (77.6 to 92.7) | 35.6 (23.3 to 50.1) | 7.6 (2.7 to 19.3) | 1.8 |
| Habiganj | No | 91 | 9.3 (3.6 to 22.0) | 0 (0 to 4.0) | 0 (0 to 4.0) | n/a |
| Jamalpur | No | 92 | 14.0 (7.7 to 24.2) | 0 (0 to 3.9) | 0 (0 to 3.9) | n/a |
| Jashore | Yes | 93 | 53.9 (41.9 to 65.5) | 7.1 (3.2 to 15.4) | 0 (0 to 3.9) | 1.1 |
| Jhalokati | Yes | 92 | 99.0 (93.1 to 99.9) | 29.9 (19.0 to 43.8) | 7.5 (2.8 to 18.9) | 1.7 |
| Jhenaidah | No | 92 | 36.6 (25.0 to 50.0) | 0.9 (0.1 to 6.2) | 0 (0 to 3.9) | 0.8 |
| Joypurhat | No | 94 | 6.2 (1.6 to 21.1) | 0 (0 to 3.8) | 0 (0 to 3.8) | n/a |
| Khagrachhari | No | 88 | 3.6 (1.1 to 11.0) | 1.1 (0.1 to 7.8) | 0 (0 to 4.1) | 1.0 |
| Khulna | Yes | 78 | 59.8 (46.9 to 71.4) | 27.0 (17.7 to 38.9) | 10.5 (4.8 to 21.5) | 1.1 |
| Kishoreganj | No | 88 | 15.7 (8.2 to 28.0) | 0 (0 to 4.1) | 0 (0 to 4.1) | n/a |
| Kurigram | No | 92 | 24.7 (16.1 to 36.0) | 3.7 (1.2 to 11.0) | 0 (0 to 3.9) | 1.1 |
| Kushtia | No | 91 | 69.1 (55.7 to 80.0) | 0 (0 to 4.0) | 0 (0 to 4.0) | n/a |
| Lakshmipur | Yes | 90 | 40.5 (28.3 to 53.9) | 19.9 (11.6 to 31.9) | 4.3 (1.1 to 15.9) | 1.4 |
| Lalmonirhat | No | 91 | 1.0 (0.1 to 7.1) | 0 (0 to 4.0) | 0 (0 to 4.0) | n/a |
| Madaripur | No | 89 | 87.4 (78.2 to 93.1) | 40.3 (27.8 to 54.1) | 2.9 (0.9 to 9.1) | 1.6 |
| Magura | No | 92 | 40.5 (28.6 to 53.5) | 0 (0 to 3.9) | 0 (0 to 3.9) | n/a |
| Manikganj | No | 90 | 45.8 (33.6 to 58.6) | 0 (0 to 4.0) | 0 (0 to 4.0) | n/a |
| Meherpur | No | 90 | 78.7 (67.3 to 87.0) | 10.0 (4.9 to 19.5) | 0 (0 to 4.0) | 1.2 |
| Moulvibazar | No | 91 | 0 (0 to 4.0) | 0 (0 to 4.0) | 0 (0 to 4.0) | n/a |
| Munshiganj | No | 90 | 52.0 (37.7 to 65.9) | 14.0 (7.4 to 24.9) | 1.1 (0.1 to 7.6) | 1.3 |
| Mymensingh | No | 85 | 6.8 (2.6 to 16.6) | 3.1 (1.0 to 9.3) | 2.1 (0.5 to 8.7) | 0.8 |
| Naogaon | No | 91 | 11.7 (6.2 to 21.1) | 0 (0 to 4.0) | 0 (0 to 4.0) | n/a |
| Narail | Yes | 90 | 90.4 (79.3 to 95.8) | 46.3 (32.3 to 60.9) | 7.8 (3.1 to 18.1) | 1.9 |
| Narayanganj | No | 88 | 38.1 (26.2 to 51.7) | 1.1 (0.1 to 7.7) | 0 (0 to 4.1) | 1.0 |
| Narsingdi | No | 92 | 17.1 (10.1 to 27.3) | 1.3 (0.2 to 9.3) | 0 (0 to 3.9) | 1.2 |
| Natore | No | 92 | 53.6 (42.3 to 64.5) | 0 (0 to 3.9) | 0 (0 to 3.9) | n/a |
| Netrokona | No | 89 | 16.4 (8.2 to 29.9) | 1.2 (0.2 to 8.7) | 0 (0 to 4.1) | 1.1 |
| Nilphamari | No | 90 | 5.1 (1.5 to 16.0) | 0 (0 to 4.0) | 0 (0 to 4.0) | n/a |
| Noakhali | Yes | 94 | 53.5 (42.0 to 64.6) | 19.2 (10.7 to 32.1) | 5.4 (1.5 to 18.1) | 1.7 |
| Pabna | No | 93 | 25.3 (16.7 to 36.4) | 1.1 (0.1 to 7.7) | 0 (0 to 3.9) | 1.0 |
| Panchagarh | No | 93 | 7.1 (3.0 to 15.8) | 4.8 (2.0 to 11.4) | 4.8 (2.0 to 11.4) | 0.9 |
| Patuakhali | Yes | 92 | 61.9 (46.9 to 74.9) | 5.5 (1.9 to 14.7) | 1.1 (0.1 to 7.6) | 1.4 |
| Pirojpur | Yes | 83 | 79.8 (67.0 to 88.5) | 48.5 (34.8 to 62.3) | 15.8 (8.9 to 26.3) | 1.6 |
| Rajbari | No | 91 | 80.9 (69.7 to 88.6) | 0 (0 to 4.0) | 0 (0 to 4.0) | n/a |
| Rajshahi | No | 93 | 40.2 (28.4 to 53.2) | 1.2 (0.2 to 8.3) | 0 (0 to 3.9) | 1.1 |
| Rangamati | No | 90 | 0 (0 to 4.0) | 0 (0 to 4.0) | 0 (0 to 4.0) | n/a |
| Rangpur | No | 91 | 5.4 (2.2 to 12.7) | 0 (0 to 4.0) | 0 (0 to 4.0) | n/a |
| Satkhira | Yes | 77 | 62.7 (48.0 to 75.4) | 10.8 (4.9 to 22.0) | 4.9 (1.6 to 14.4) | 1.3 |
| Shariatpur | Yes | 91 | 73.9 (63.0 to 82.5) | 4.9 (1.8 to 12.6) | 0 (0 to 4.0) | 1.1 |
| Sherpur | No | 92 | 3.7 (1.2 to 11.3) | 0 (0 to 3.9) | 0 (0 to 3.9) | n/a |
| Sirajganj | No | 90 | 12.3 (6.6 to 21.7) | 2.0 (0.5 to 7.9) | 0 (0 to 4.0) | 0.9 |
| Sunamganj | No | 89 | 13.6 (6.4 to 26.7) | 0 (0 to 4.1) | 0 (0 to 4.1) | n/a |
| Sylhet | No | 92 | 0 (0 to 3.9) | 0 (0 to 3.9) | 0 (0 to 3.9) | n/a |
| Tangail | No | 94 | 14.2 (7.5 to 25.2) | 0 (0 to 3.8) | 0 (0 to 3.8) | n/a |
| Thakurgaon | No | 92 | 1.2 (0.2 to 8.6) | 0 (0 to 3.9) | 0 (0 to 3.9) | n/a |

*Household-weighted percentages with logit 95% confidence intervals. The design effect is the design-based variance over the variance under simple random sampling of the same size, including weighting; n/a where no household exceeded the cut-off. District names follow current official spelling. With a median of 91 households per district, the median interval at 1,500 µS/cm spans 12 percentage points among the 41 districts with any household above 1,500 µS/cm (8 points across all 64), and at 700 µS/cm 20 points. Panchagarh’s shares above 1,500 and 3,000 µS/cm come entirely from five readings that probably reflect recording errors, and Gaibandha’s share above 1,500 µS/cm includes a sixth such reading (main text; S3 Table). These estimates should not be used to rank districts.*
