## Supplementary material for "Source-water salinity measured by electrical conductivity in Bangladesh, 2025: a nationally representative cross-sectional survey": S5_Table.docx

### S5 Table. Numeric data for the figures and text

Not tabulated here: S1 Fig (conceptual) and Fig 2A (interpolated surface of 3,124 cluster means). Figs 2C and 2D are in S4 Table, Panel A. Fig 4B is in S6 Table, Panels A and C. Sensitivity of the 1,500 µS/cm estimate is in S3 Table. S2 Fig is summarised in S3 Table. Estimates in S3 Fig are taken from the cited studies. Panel F supports the Results text.

**Panel A. Fig 1: share of the population above each EC value, % (95% CI)**

| EC value (µS/cm) | Bangladesh | 19 coastal-zone districts | Other 45 districts |
| --- | --- | --- | --- |
| 100 | 95.3 (94.6 to 96.0) | 93.1 (91.3 to 94.5) | 96.2 (95.4 to 96.8) |
| 111 | 94.5 (93.8 to 95.2) | 92.5 (90.6 to 94.0) | 95.3 (94.5 to 96.1) |
| 123 | 93.4 (92.6 to 94.2) | 91.8 (89.9 to 93.4) | 94.1 (93.1 to 94.9) |
| 137 | 91.7 (90.8 to 92.6) | 91.0 (89.0 to 92.6) | 92.0 (90.9 to 93.0) |
| 152 | 89.9 (88.8 to 90.8) | 89.7 (87.4 to 91.5) | 89.9 (88.7 to 91.0) |
| 169 | 88.4 (87.3 to 89.4) | 89.1 (86.8 to 91.0) | 88.2 (86.9 to 89.3) |
| 187 | 86.5 (85.4 to 87.6) | 88.2 (85.8 to 90.2) | 85.9 (84.5 to 87.2) |
| 208 | 84.3 (83.0 to 85.4) | 86.8 (84.4 to 88.9) | 83.3 (81.9 to 84.7) |
| 231 | 81.8 (80.5 to 83.0) | 85.7 (83.2 to 87.9) | 80.3 (78.7 to 81.8) |
| 257 | 78.3 (76.8 to 79.6) | 84.0 (81.3 to 86.3) | 76.1 (74.4 to 77.7) |
| 285 | 74.6 (73.0 to 76.0) | 82.5 (79.7 to 84.9) | 71.6 (69.7 to 73.4) |
| 316 | 70.3 (68.7 to 71.8) | 81.2 (78.4 to 83.7) | 66.1 (64.2 to 68.0) |
| 351 | 64.4 (62.8 to 66.0) | 78.6 (75.7 to 81.2) | 59.1 (57.1 to 61.0) |
| 390 | 58.5 (56.9 to 60.1) | 75.6 (72.7 to 78.4) | 52.0 (50.1 to 53.9) |
| 433 | 53.5 (51.9 to 55.1) | 71.8 (68.9 to 74.5) | 46.6 (44.7 to 48.5) |
| 481 | 48.2 (46.6 to 49.8) | 68.5 (65.6 to 71.3) | 40.5 (38.6 to 42.4) |
| 534 | 43.4 (41.8 to 45.0) | 64.2 (61.1 to 67.1) | 35.5 (33.7 to 37.3) |
| 593 | 37.9 (36.4 to 39.5) | 59.0 (55.9 to 62.0) | 29.9 (28.3 to 31.7) |
| 658 | 31.7 (30.2 to 33.1) | 53.1 (50.2 to 56.1) | 23.5 (22.0 to 25.1) |
| 731 | 26.1 (24.7 to 27.5) | 45.4 (42.4 to 48.4) | 18.7 (17.3 to 20.3) |
| 811 | 20.5 (19.3 to 21.8) | 38.3 (35.4 to 41.3) | 13.8 (12.5 to 15.1) |
| 901 | 15.7 (14.6 to 16.9) | 32.6 (29.8 to 35.5) | 9.3 (8.3 to 10.5) |
| 1,000 | 11.8 (10.9 to 12.8) | 26.8 (24.3 to 29.5) | 6.1 (5.3 to 7.1) |
| 1,110 | 9.6 (8.8 to 10.5) | 22.9 (20.5 to 25.4) | 4.6 (3.9 to 5.4) |
| 1,233 | 7.5 (6.8 to 8.3) | 18.5 (16.4 to 20.9) | 3.3 (2.7 to 4.0) |
| 1,369 | 6.3 (5.6 to 7.0) | 16.0 (14.1 to 18.2) | 2.6 (2.1 to 3.2) |
| 1,520 | 5.0 (4.4 to 5.7) | 13.0 (11.2 to 14.9) | 2.0 (1.6 to 2.6) |
| 1,688 | 4.0 (3.5 to 4.6) | 10.7 (9.2 to 12.5) | 1.5 (1.1 to 2.0) |
| 1,874 | 3.3 (2.8 to 3.9) | 9.1 (7.7 to 10.7) | 1.1 (0.8 to 1.6) |
| 2,081 | 2.6 (2.2 to 3.1) | 7.3 (6.0 to 8.8) | 0.9 (0.6 to 1.3) |
| 2,310 | 2.1 (1.7 to 2.5) | 5.8 (4.7 to 7.3) | 0.6 (0.4 to 1.0) |
| 2,565 | 1.5 (1.2 to 2.0) | 4.6 (3.5 to 5.9) | 0.4 (0.2 to 0.7) |
| 2,848 | 1.2 (0.9 to 1.6) | 3.4 (2.5 to 4.5) | 0.4 (0.2 to 0.7) |
| 3,162 | 1.0 (0.7 to 1.4) | 2.7 (1.9 to 3.9) | 0.3 (0.2 to 0.6) |
| 3,511 | 0.8 (0.5 to 1.1) | 2.0 (1.3 to 3.1) | 0.3 (0.2 to 0.6) |
| 3,899 | 0.6 (0.4 to 0.9) | 1.5 (0.9 to 2.5) | 0.2 (0.1 to 0.5) |
| 4,329 | 0.4 (0.2 to 0.7) | 1.0 (0.5 to 1.7) | 0.2 (0.1 to 0.4) |
| 4,806 | 0.2 (0.1 to 0.4) | 0.5 (0.2 to 1.1) | 0.1 (0.1 to 0.3) |
| 5,337 | 0.2 (0.1 to 0.4) | 0.4 (0.2 to 1.1) | 0.1 (0.1 to 0.3) |
| 5,926 | 0.2 (0.1 to 0.4) | 0.3 (0.1 to 1.0) | 0.1 (0.1 to 0.3) |
| 6,579 | 0.1 (0.1 to 0.3) | 0.2 (0.0 to 1.0) | 0.1 (0.1 to 0.3) |
| 7,305 | 0.1 (0.0 to 0.3) | 0.0 (0.0 to 0.2) | 0.1 (0.1 to 0.3) |
| 8,111 | 0.1 (0.0 to 0.2) | 0.0 (0.0 to 0.1) | 0.1 (0.0 to 0.3) |
| 9,006 | 0.0 (0.0 to 0.1) | 0.0 (0.0 to 0.1) | 0.1 (0.0 to 0.1) |
| 10,000 | 0.0 (0.0 to 0.0) | 0.0 (0.0 to 0.1) | 0.0 (0.0 to 0.1) |

**Panel B. Fig 2B: cluster median EC by elevation band**

| Area | Elevation band (m) | Clusters (n) | Median EC, µS/cm (interquartile range) |
| --- | --- | --- | --- |
| Coastal zone | 0 to 5 | 250 | 826 (578 to 1,236) |
| Coastal zone | 5 to 10 | 585 | 890 (624 to 1,294) |
| Coastal zone | 10 to 20 | 66 | 296 (153 to 513) |
| Coastal zone | 20 to 50 | 19 | 227 (136 to 394) |
| Coastal zone | above 50 | 1 (not plotted) | 626 (626 to 626) |
| Other districts | 0 to 5 | 21 | 677 (593 to 1,238) |
| Other districts | 5 to 10 | 525 | 630 (442 to 852) |
| Other districts | 10 to 20 | 941 | 482 (316 to 724) |
| Other districts | 20 to 50 | 510 | 312 (194 to 486) |
| Other districts | above 50 | 206 | 188 (119 to 287) |

**Panel C. Fig 3A: households above 1,500 µS/cm (national and zone estimates are in Table 2 and S4 Table, Panel A)**

| Block | Group | Households (n) | Households above 1,500, % (95% CI) |
| --- | --- | --- | --- |
| Residence | Rural | 4,403 | 5.7 (5.0 to 6.5) |
| Residence | Urban | 1,637 | 4.4 (3.5 to 5.6) |
| Wealth (poorest to richest) | Quintile 1 | 1,476 | 4.5 (3.5 to 5.8) |
| Wealth (poorest to richest) | Quintile 2 | 1,296 | 6.8 (5.6 to 8.3) |
| Wealth (poorest to richest) | Quintile 3 | 1,192 | 5.6 (4.3 to 7.2) |
| Wealth (poorest to richest) | Quintile 4 | 1,098 | 4.7 (3.6 to 6.1) |
| Wealth (poorest to richest) | Quintile 5 | 978 | 4.8 (3.7 to 6.3) |
| Household | Child under 5 | 2,058 | 5.3 (4.3 to 6.5) |
| Household | No child under 5 | 3,982 | 5.3 (4.6 to 6.1) |
| Household | No pregnant woman | 4,859 | 5.3 (4.7 to 6.1) |
| Household | Pregnant woman | 284 | 5.5 (3.5 to 8.5) |
| Tested source | Deep tube well | 1,458 | 11.9 (10.2 to 13.7) |
| Tested source | Other | 167 | 2.5 (0.9 to 7.0) |
| Tested source | Piped | 979 | 2.7 (1.7 to 4.1) |
| Tested source | Shallow tube well | 3,436 | 4.2 (3.4 to 5.0) |

**Panel D. Fig 3B: main source not in the dwelling or yard**

| Area | EC group | Households (n) | Source not in dwelling or yard, % (95% CI) |
| --- | --- | --- | --- |
| Bangladesh | EC above 1,500 | 432 | 36.9 (31.7 to 42.4) |
| Bangladesh | EC up to 1,500 | 5,592 | 13.8 (12.8 to 14.8) |
| Coastal zone | EC above 1,500 | 319 | 44.5 (38.2 to 51.0) |
| Coastal zone | EC up to 1,500 | 1,443 | 31.6 (28.9 to 34.4) |

**Panel E. Fig 4A: people in households with EC above 1,500 µS/cm, by arsenic**

| Area | Category | Members, % (95% CI) | People, millions (95% CI) |
| --- | --- | --- | --- |
| Bangladesh | EC above 1,500, arsenic at or below 10 µg/L | 4.0 (3.5 to 4.5) | 6.7 (5.9 to 7.7) |
| Bangladesh | EC above 1,500 and arsenic above 10 µg/L | 1.3 (1.0 to 1.7) | 2.2 (1.7 to 3.0) |
| 19 coastal-zone districts | EC above 1,500, arsenic at or below 10 µg/L | 10.1 (8.6 to 11.8) | 4.5 (3.9 to 5.3) |
| 19 coastal-zone districts | EC above 1,500 and arsenic above 10 µg/L | 3.7 (2.7 to 5.0) | 1.7 (1.2 to 2.3) |
| Other 45 districts | EC above 1,500, arsenic at or below 10 µg/L | 1.6 (1.2 to 2.2) | 2.0 (1.6 to 2.7) |
| Other 45 districts | EC above 1,500 and arsenic above 10 µg/L | 0.4 (0.2 to 0.7) | 0.5 (0.3 to 0.9) |

**Panel F. Population by EC band, % (Results)**

| Area | ≤700 | >700 to 1,000 | >1,000 to 1,500 | >1,500 to 3,000 | >3,000 |
| --- | --- | --- | --- | --- | --- |
| Bangladesh | 71.6 | 16.6 | 6.5 | 4.2 | 1.1 |
| Coastal zone | 51.5 | 21.7 | 13.0 | 10.8 | 3.0 |
| Other districts | 79.2 | 14.6 | 4.1 | 1.7 | 0.3 |

**Panel G. Weighted median EC by month (exploratory)**

| Area | Month | Households (n) | Median EC, µS/cm (interquartile range) |
| --- | --- | --- | --- |
| Coastal zone | February | 87 | 594 (405 to 826) |
| Coastal zone | March | 507 | 653 (392 to 948) |
| Coastal zone | April | 492 | 670 (404 to 1,098) |
| Coastal zone | May | 475 | 762 (403 to 1,216) |
| Coastal zone | June | 210 | 777 (400 to 1,210) |
| Other districts | February | 218 | 395 (281 to 610) |
| Other districts | March | 1,255 | 403 (286 to 625) |
| Other districts | April | 1,102 | 412 (263 to 645) |
| Other districts | May | 1,259 | 410 (253 to 658) |
| Other districts | June | 433 | 439 (279 to 681) |

**Panel H. Other estimates cited in the text**

| Measure | Estimate (95% CI) |
| --- | --- |
| Counts scaled to the 2025 testing period (factor 1.034): above 1,500 / 3,000 / 1,000 µS/cm | 9.3 / 1.9 / 20.8 million |
| Share of households above 1,500 / 3,000 µS/cm located in the coastal zone | 70.4% / 76.9% |
| Moran’s I of cluster-mean log10 EC: raw; after residence, wealth, elevation, rainfall and division; plus spatial smooth | 0.585 (P = 0.001); 0.349; 0.13 |
| Main source not in the dwelling or yard, coastal zone, EC above 1,500 against up to 1,500 µS/cm: adjusted prevalence ratio (division; district) | 1.27 (1.08 to 1.49); 1.19 (0.99 to 1.43) |
| Deep against shallow tube wells, national prevalence ratio at 1,500 µS/cm: crude; district | 2.85 (2.24 to 3.62); 0.79 (0.64 to 0.99) |
| Weighted mean TDS equivalent (0.64 x EC), Khulna and Barishal: shallow (n = 570); deep (n = 695) tube wells | 619 mg/L (572 to 665); 744 mg/L (693 to 795) |
| EC above 1,500 µS/cm, prevalence ratio per wealth quintile (adjusted for residence, tested source and division): national without Sylhet; coastal zone | 1.02 (0.93 to 1.11); 0.99 (0.90 to 1.09) |
| EC above 1,500 µS/cm, urban against rural (adjusted for wealth quintile, tested source and division) | 0.90 (0.66 to 1.23) |
| People in households above the ECR TDS limit of 1,000 mg/L (above 1,560 µS/cm), 1997 and 2023 Rules | 4.7% (4.1 to 5.3) |
| People whose source had arsenic at or below 10 µg/L, in households above 1,500 µS/cm: national; coastal zone | 4.7% (4.1 to 5.3); 12.3% (10.5 to 14.4) |
| Tested tube wells located in the coastal zone (household weighted): deep; shallow | 59.9% (57.1 to 62.6); 14.5% (13.4 to 15.7) |
| Coastal households, EC above 1,500 against at or below 1,500 µS/cm: a different source used in the wet season | 5.9% (3.3 to 10.3) against 3.4% (2.4 to 4.8) |
| Coastal households that treat their drinking water | 9.8% (8.3 to 11.6) |
| Khulna division share of coastal tests above zero, February to June (unweighted; n = 87, 491, 491, 468, 209) | 16.1%, 15.1%, 17.3%, 33.1%, 41.1% |

*Percentages are household weighted except Panels A, E and F and the population rows of Panel H, which are member weighted. People = member-weighted proportion x reference population (169.8 million; 45.0 million for the coastal zone; 124.8 million for the other districts). Clusters with fewer than 10 per band are not plotted in Fig 2B.*
