## Supplementary material for "Source-water salinity measured by electrical conductivity in Bangladesh, 2025: a nationally representative cross-sectional survey": S6_Table.docx

### S6 Table. Sensitivity of the depth and month comparisons to the specification

Geometric mean ratios (GMR) from survey-weighted linear models on log10 EC, with 54 zero readings excluded. Source type is the tested source. FE, fixed effects. **Declared primary specifications are in bold.**

**Panel A. Depth: deep against shallow tube wells**

| Specification | Coastal zone (n = 1,433) | Other districts (n = 3,435) |
| --- | --- | --- |
| Crude | 1.09 (0.96 to 1.23) | 1.07 (0.99 to 1.16) |
| Residence, wealth, division | 0.86 (0.75 to 0.98) | 1.20 (1.10 to 1.31) |
| Residence, wealth, district | 0.82 (0.73 to 0.93) | 1.06 (0.97 to 1.15) |
| **District + elevation + rainfall + residence + wealth (primary)** | **0.82 (0.73 to 0.92)** | **1.04 (0.96 to 1.13)** |
| Primary, by coastal region (interaction P < 0.001): Khulna and Barishal (n = 826) | 0.86 (0.70 to 1.06) | not applicable |
| Primary, by coastal region: Chattogram (n = 432) | 0.70 (0.59 to 0.83) | not applicable |
| Primary, by coastal region: Gopalganj and Shariatpur, Dhaka division (n = 175) | 1.43 (1.17 to 1.75) | not applicable |
| Primary + measurer FE | 0.83 (0.73 to 0.93) | 1.05 (0.97 to 1.13) |
| Upazila FE + elevation + rainfall + residence + wealth | 0.93 (0.80 to 1.07) | 1.02 (0.95 to 1.10) |
| Crude, in the 851 households of the upazila model | 0.97 (0.84 to 1.12) | not applicable |
| Primary, in the 851 households of the upazila model | 0.88 (0.77 to 0.99) | not applicable |

*In the coastal zone, 69 of 142 tube-well upazilas (851 of 1,433 households) had both well types.*

**Panel B. Month: May-June and April against February-March**

| Sample and specification | n | GMR May-June (95% CI) | GMR April (95% CI) |
| --- | --- | --- | --- |
| Coastal zone, district FE + residence + wealth | 1,746 | 1.30 (1.08 to 1.56) | 1.05 (0.91 to 1.22) |
| **Coastal zone, district FE + source + elevation + rainfall + residence + wealth** | 1,746 | **1.17 (0.98 to 1.39)** | 0.93 (0.82 to 1.06) |
| Coastal zone, primary + measurer FE | 1,746 | 1.24 (0.95 to 1.61) | 0.91 (0.80 to 1.04) |
| Coastal zone, mixed model (unweighted), district and measurer random intercepts | 1,746 | 1.21 (1.05 to 1.40) | 0.94 (0.85 to 1.04) |
| Coastal zone, upazila FE + source + elevation + rainfall + residence + wealth | 1,746 | 0.86 (0.61 to 1.21) | 0.79 (0.59 to 1.06) |
| Coastal deep tube wells, primary without source | 912 | 0.99 (0.87 to 1.14) | 1.06 (0.94 to 1.18) |
| Coastal shallow tube wells, primary without source | 521 | 1.27 (0.82 to 1.97) | 0.92 (0.70 to 1.22) |
| Coastal piped water, primary without source | 221 | 1.37 (0.86 to 2.19) | 0.71 (0.45 to 1.11) |
| Other districts, primary | 4,238 | 0.94 (0.87 to 1.01) | 0.97 (0.91 to 1.03) |
| Other districts, upazila FE | 4,238 | 1.02 (0.86 to 1.20) | 0.92 (0.79 to 1.06) |

*Upazila fixed effects are identified only from the 31 of 156 coastal upazilas with tests in at least two month groups (471 of 1,746 households). Weighted median EC by month is in S5 Table, Panel G.*

**Panel C. Excluding all readings of 20 µS/cm or less**

| Depth: deep against shallow tube wells | Coastal zone (n = 1,422) | Other districts (n = 3,410) |
| --- | --- | --- |
| Crude | 1.04 (0.94 to 1.16) | 1.06 (0.99 to 1.14) |
| Residence, wealth, division | 0.82 (0.74 to 0.91) | 1.19 (1.09 to 1.29) |
| Residence, wealth, district | 0.78 (0.71 to 0.86) | 1.03 (0.96 to 1.12) |
| **District + elevation + rainfall + residence + wealth (primary)** | **0.78 (0.72 to 0.86)** | **1.02 (0.94 to 1.10)** |
| Primary + measurer FE | 0.79 (0.72 to 0.86) | 1.03 (0.96 to 1.10) |
| Upazila FE + elevation + rainfall + residence + wealth | 0.88 (0.80 to 0.97) | 1.00 (0.94 to 1.06) |
| Primary, in the 845 households of the upazila model | 0.84 (0.77 to 0.93) | not applicable |

| Month: against February-March | n | GMR May-June (95% CI) | GMR April (95% CI) |
| --- | --- | --- | --- |
| Coastal zone, primary | 1,723 | 1.23 (1.05 to 1.44) | 1.00 (0.90 to 1.13) |
| Coastal zone, primary + measurer FE | 1,723 | 1.36 (1.05 to 1.75) | 0.98 (0.87 to 1.11) |
| Coastal zone, upazila FE | 1,723 | 0.97 (0.72 to 1.31) | 0.88 (0.67 to 1.16) |
| Other districts, primary | 4,209 | 0.95 (0.88 to 1.02) | 1.00 (0.95 to 1.05) |
| Other districts, primary + measurer FE | 4,209 | 1.06 (0.96 to 1.17) | 1.02 (0.96 to 1.07) |
| Other districts, upazila FE | 4,209 | 1.03 (0.91 to 1.15) | 0.98 (0.89 to 1.09) |

*Same specifications as Panels A and B with readings of 20 µS/cm or less excluded (11 coastal tube-well households and 23 coastal households in the month sample). The 106 excluded readings are described in S3 Table.*
